# RNA-sequencing improves diagnosis for neurodevelopmental disorders by identifying pathogenic non-coding variants and reinterpretation of coding variants

**DOI:** 10.1101/2022.06.05.22275956

**Authors:** Jordy Dekker, Rachel Schot, Michiel Bongaerts, Walter G. de Valk, Monique M. van Veghel-Plandsoen, Kathryn Monfils, Hannie Douben, Peter Elfferich, Esmee Kasteleijn, Leontine M.A. van Unen, Geert Geeven, Jasper J. Saris, Yvette van Ierland, Frans W. Verheijen, Marianne L.T. van der Sterre, Farah Sadeghi Niaraki, Hidde H. Huidekoper, Monique Williams, Martina Wilke, Virginie J.M. Verhoeven, Marieke Joosten, Anneke J.A. Kievit, Ingrid M.B.H. van de Laar, Lies H. Hoefsloot, Marianne Hoogeveen-Westerveld, Mark Nellist, Grazia M.S. Mancini, Tjakko J. van Ham

## Abstract

**Background:** For neurodevelopmental disorders (NDD), a molecular diagnosis is key for predicting outcome, treatment and genetic counseling. Currently, in about half of NDD cases, routine DNA-based testing fails to establish a genetic diagnosis. Transcriptome analysis (RNA-seq) improves the diagnostic yield for some groups of diseases, but has not been applied to NDD in a routine diagnostic setting.

**Methods:** Here, we explored the diagnostic potential of RNA-seq in a cohort of 96 individuals including 67 undiagnosed NDD subjects. We created a user-friendly web-application to analyze RNA-seq data from single individuals’ cultured skin fibroblasts for genic, exonic and intronic expression outliers, based on modified OUTRIDER Z-scores. Candidate pathogenic events were complemented/matched with genomic data and, if required, confirmed with additional functional assays.

**Results:** We identified pathogenic small genomic deletions, mono-allelic expression, aberrant splicing events, deep intronic variants resulting in pseudo-exon insertion, but also synonymous and nonsynonymous variants with deleterious effects on transcription. This approach increased the diagnostic yield for NDD by 12%. Diagnostic pitfalls during transcriptome analysis include detection of splice abnormalities in putative disease genes caused by benign polymorphisms and/or absence of expression of the responsible gene in the tissue of choice. This was misleading in one case and could have led to the wrong diagnosis in the absence of appropriate phenotyping.

**Conclusions:** Nonetheless, our results demonstrate the utility of RNA-seq in molecular diagnostics and stress the importance of multidisciplinary team consultation. In particular, the approach is useful for the identification and interpretation of unexpected pathogenic changes in mRNA processing and expression in NDD.

## Background

Neurodevelopmental disorders (NDD) involve an impaired development of the central nervous system often resulting in intellectual disability (ID) and other chronic neurological or behavioral problems like autism, motor impairment and epilepsy. The majority of NDD are thought to have an underlying genetic cause (1). In the last decade, the implementation of high throughput sequencing techniques, e.g. whole exome sequencing (WES), has helped increase the diagnostic yield in NDD to approximately 36% (2).

One drawback of WES analysis algorithms is the inability to interpret the potential pathogenic effects of variants that could affect pre-mRNA splicing or gene expression. Furthermore, it is estimated that for Mendelian disease up to one third of the pathogenic variants are located in non-coding regions, which are not covered by clinical WES (3). In diagnostic settings, synonymous single nucleotide variants (SNVs) are often classified as benign, despite numerous examples where synonymous SNVs affect mRNA splicing and stability and cause disease (4, 5). Whole genome sequencing (WGS) improves the detection of non-coding variants (6). However, in addition to the high costs of WGS, functional annotation for the vast majority of these non-coding variants is currently lacking, making interpretation of pathogenicity very difficult.

To increase the diagnostic yield in individuals with a genetic disorder, RNA-sequencing (RNA-seq) can be a complementary tool to DNA-based analysis, as it detects the effects of both coding and non-coding genetic variants on RNA transcripts (7–9). In addition to relative over- or under-expression, aberrant splicing and skewed or mono-allelic expression (MAE) can be identified (8, 9). Recently, it was estimated that ∼30% of variants of unknown significance (VUSs) could be reclassified as either likely benign or likely pathogenic after performing RNA-seq analysis (10). Several RNA-seq studies identified disease-causing variants in WES-negative cases, with a diagnostic rate between 7.5% and 36%, focusing on individuals with a muscle disorder (8), neuromuscular disorders (11), mitochondrial disease (9, 12) or in a heterogeneous disease population (13–16), indicating that RNA-seq is an effective complementary tool to standard genetic diagnostics. However, previous RNA-seq studies either assumed *a priori* knowledge of DNA variants, focused on consanguineous populations, employed not routinely applicable bioinformatics pipelines, or used RNA from tissues that are not easily accessible, preventing widespread and routine clinical application. Here we apply a newly developed diagnostic flow for RNA-seq-based analysis of skin fibroblast RNA, including a user-friendly analysis tool, in a selected population of undiagnosed NDD individuals. We identified causative pathogenic variants in 8/67 undiagnosed NDDs (12%), including small genomic deletions, variants causing MAE, and variants that affect splicing, proving potential of this approach in the diagnostic setting.

## Methods

### Patient cohort, genome sequencing, gene panels and selection for RNA-seq

All individuals were evaluated at the Department of Clinical Genetics, Erasmus Medical Center, Rotterdam, the Netherlands and examined by a clinical geneticist who provided pre-test counseling. Informed consent for all diagnostic tests and publication was obtained (IRB protocol METC-2012-387). Diagnostic genomic microarrays and (neonatal) metabolic screening preceded Next Generation Sequencing (NGS) tests. Diagnostic whole exome sequencing (WES) was performed, with analysis of gene panels for intellectual disability (ID, trio analysis, i.e. including parental DNA), brain malformations (NEUMIG, single) or epilepsy (EPI, single), autism (AUT, single) or Ciliopathy (single) followed by full trio exome analysis of *de novo*, recessive and X-linked variants, as described previously (17). Furthermore, multiplex ligation-dependent probe amplification (MLPA) analysis of known recurrent deletion loci (e.g. *LIS1*) and repeat areas (e.g. *ARX*) was performed, covering variants not detected by WES. The composition of the gene panels used for WES and RNA-seq analysis can be found in **Table S1** or at (18) (panel composition is updated twice per year). Variant classification was performed according to international criteria (19). Variant annotation was done according to the HGVS-nomenclature guidelines (20). Alamut Visual v2.15 was used for splice predictions. Individuals with negative results (either no candidate pathogenic variant, or variants of unknown significance) were selected for further investigation during a bi-weekly multidisciplinary team meeting at the Department of Clinical Genetics, with the participation of laboratory specialists, neurologists, clinical geneticists and basic research scientists. Prioritization criteria were the presence of a skin biopsy previously sampled for diagnostic purposes and the clinical suspicion of a genetic cause based on disease course, on the presence of associated anomalies/dysmorphism or familial occurrence, age and relevance for reproductive risk. After the RNA-seq analysis, results were communicated and the families were counseled by a clinical geneticist.

### Cell culture and cycloheximide treatment

Skin biopsy-derived fibroblasts were grown in Ham’s F10 medium containing 15% (v/v) fetal calf serum (FCS) and 1% (v/v) penicillin/streptomycin (PS), according to standard procedures in the ISO certified laboratory of the Clinical Genetics department, including mycoplasma testing (21). For isolation of total RNA, cells were cultured to 70-80% confluency in 75 cm^2^ tissue culture flasks. Cycloheximide (CHX) (Sigma-Aldrich) treatment was 100 µg/ml for 24h.

### RNA-sequencing, read alignment and quality control

The RNeasy mini kit (Qiagen, Venlo, The Netherlands) was used for RNA isolation according to the manufacturer’s protocol. Quality of RNA was assessed by determining the RNA integrity number (RIN) using a Bioanalyzer (Agilent); fibroblast RNA typically had a RIN ≥ 10. Next, polyadenylated RNA (mRNA) transcripts were enriched using the NEBNext PolyA kit (NEB), followed by fragmentation and cDNA synthesis using the NEBNext RNA Ultra II Directional kit (NEB). Strand specific sequencing (polyA enriched and Globin reduction for PAX samples) of total RNA samples was performed (GenomeScan, Leiden, The Netherlands) on an Illumina NovaSeq 6000 with 150 bp Paired-end reads including unique molecular identifier (UMI)-adaptors. A minimum of 40 million reads were generated per sample. FASTQ datasets were processed using an established pipeline at the Department of Clinical Genetics (Erasmus MC, Rotterdam, the Netherlands). Reads were mapped using HiSat2 (v2.2.1) and Kallisto (v0.44.0) software to the hg19 GENCODE genome assembly (22, 23). Alignment of sequence reads to SAM format was done by HiSat2. SAM to BAM conversion and marking of duplicate reads was done by Samtools (v1.9), followed by Bedtools (v2.29.2) to count the reads per coding sequence (CDS). Uniquely mapped reads with a MAPQ score < 1 were removed (24). Quality control was performed using RNA-SeQC (v4.0.0) and Samtools flagstat (v1.9) with typical *in silico* quality parameters for fragment size: mean inner distance > 20, inner distance peak ratio >1.35, percentage mapped reads > 85 %, percentage duplicate reads < 50% and Transcript integrity number (TIN) values > 70 (24, 25). Variants were called using GATK4 (v4.1.9). Data was visualized in the Integrated Genome Viewer (IGV; v2.4.15) using GRCh37 – hg19 as reference (26)

### Computing differential expression: Z-scores and p-values

After quality control raw counts were normalized using the OUTRIDER (Outlier in RNA-seq finder) algorithm to compare the sample under investigation to the rest of the cohort (27). In addition to calculating Z-scores and p-values per gene, similar analyses were performed per exon and per intron, allowing the detection of aberrant expression and aberrant pre-mRNA splicing. Z-score data were made accessible via a self-developed user-friendly Jupyter-based web browser application. This allows non-bioinformatically trained genome analysts to select individualized settings including type of Z-scores (gene, exon, intron level), WES-panel or ROH to be analysed and Z-score/p-value thresholds (**Fig. 1C-5**). Data visualizations essential for analysis include PCA plot of all samples included, Z-score distribution plot, volcano plot, exon-ranking plot, fragment plot gene of interest and transcriptome-wide outlier plots indicating all Z-scores plotted along the chromosome (**Fig. 1C**). As well, HPO clinical phenotyping terms could be entered to assist in identifying specific changes in relevant genes, and lists can be generated and exported with genes/exons/intron showing altered Z-scores, and for example genes not expressed within specific exome gene panels.

**Fig. 1.**
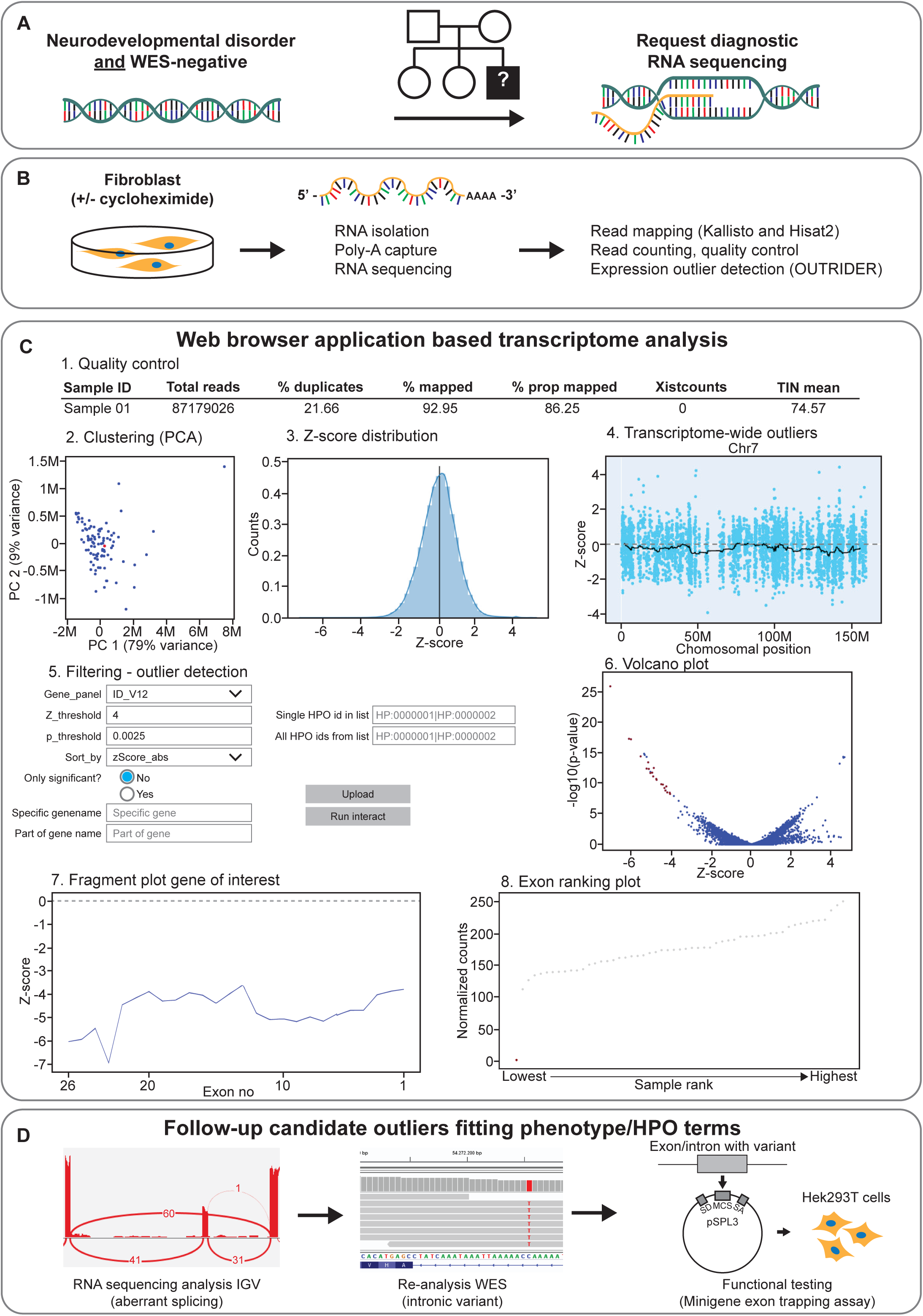
RNA-seq procedure in individuals with undiagnosed neurodevelopmental disorder (NDD). **A** 67 Individuals with a NDD for whom no molecular diagnosis was obtained by routine WES (WES-negative) were selected for diagnostic RNA-seq. **B** Total RNA was isolated from fibroblasts either treated with CHX or left untreated. Poly-A capture was performed to enrich the mRNA fraction and sequenced to a minimum of 40 million total reads. Reads were mapped using Kallisto and Hisat2 algorithms, read counts were corrected using the OUTRIDER algorithm. **C** Overview of our web browser application tool output to analyze RNA-seq data. C1: Quality control of the sample, where QC parameters that do not reach our cut-off are normally indicated in red. C2: PCA plot where the analyzed sample (red dot) is compared to all the other samples in the cohort (blue dots). C3: Z-score distribution plot at either gene, exon or intron level. C4: Z-scores along chromosome 7 of one individual; this plot can be generated for each chromosome separately. C5: Web browser application menu where you can set filters (e.g. gene panel, Z-score and p-value threshold, HPO terms, specific gene) for the analysis. C6: Volcano plot indicating the significantly under- and over-expressed genes/exons or introns within the analyzed panel (red dots). C7: Fragment plot showing all exonic Z-scores of one gene of the sample that is being analyzed. C8: Exon ranking plot showing one exon of one gene of the sample being analyzed (red dot) vs. all other samples in the cohort (grey dots). **D** After fitting the gene outlier to phenotype/HPO terms of the individual, analysis of the mapped sequences in IGV v2.4.15 is required to identify the cause of the outlier. Next, re-analysis of the WES is performed to identify the genomic variant. A functional test is often required to validate the effect seen in the RNA-seq data.

### Mono-allelic expression analysis

Genes showing MAE identified by our exonic Z-score expression outlier detection pipeline were confirmed using the DROP pipeline (28). Mapped RNA-Seq reads (HiSat2) and germline DNA variants from the Erasmus MC Clinical Genetics department routine diagnostic WES workflow (GATK HaplotypeCaller Version 3.7-0-gcfedb67) of 37 individuals’ fibroblast samples were processed with the MAE module of the DROP pipeline (v1.0.5) using GENCODE transcript annotation (v34lift37) and default configuration parameters.

### Confirmation of aberrations detected by RNA-seq

#### Genomic confirmation and minigene exon trapping assay

After detection by RNA-seq of outlier expression and candidate variants, DNA sequences were retrospectively verified in WES/WGS data and, in case of doubt by Sanger sequencing. In some cases, RT-PCR was performed according to standard procedures in order to confirm splicing aberrations (21). To confirm effects on splicing a minigene exon trapping assays were performed. The exon(s) of interest and surrounding intronic sequences were amplified from genomic DNA and cloned into the pSPL3 vector (Thermo Fisher Scientific, Invitrogen) using Gibson assembly (29). All constructs were verified by Sanger sequencing. 1 µg of the minigene construct was transfected into 70-90% confluent HEK 293T cells growing in a 12-wells plate using 5 µg polyethyleneimine. After 24 and 48 hours incubation, RNA was isolated as described above and cDNA was generated using the iScript cDNA synthesis kit (BioRad). A standard set of primers was used to amplify the products transcribed from the vector. RT-PCR products were analyzed by agarose gel electrophoresis and Sanger sequencing. Primer sequences are available on request.

#### Western blot (TBC1D7)

Fibroblasts were cultured in Dulbecco’s modified Eagle medium (DMEM) (Lonza, Verviers, Belgium) containing 10% (v/v) FCS), 1% (v/v) PS, in 10% CO2. Confluent cells were transferred to ice, washed with phosphate-buffered saline (PBS) lysed in 50mM Tris-HCl pH8.0, 150 mM NaCl, 50 mM NaF, and 1% Triton X-100 containing protease inhibitor (Complete, Roche Molecular Biochemicals, Woerden, The Netherlands). Lysates were centrifuged at 10,000 g for 10 min.at 4°C, supernatant was recovered and diluted in Laemmli sample buffer (Bio-Rad). Samples were heated at 96 °C for 5 minutes prior to loading samples on 4-15% Criterion™ TGX Stain-Free™ Protein Gel (Bio-Rad). Proteins were transferred to a nitrocellulose membrane using a Trans-Blot Turbo system at 2.5 A, 25V for 10 min. Blocking was done in 5% low-fat milk powder (Elk, Campina) for 30 minutes while shaking. After blocking the blots were washed 2 times in PBS. Antibodies were diluted in PBST (0.1% Tween in PBS). Primary antibodies used were 1:10,000 of 1895 (rabbit polyclonal against TSC2) (30), 1:5000 of 2197 (rabbit polyclonal against TSC1) (30), 1:5000 rabbit (Cell Signaling) + 1:2000, mouse anti-pan-AKT (Cell Signaling, 2920), 1:100,000 mouse anti-GAPDH (Abcam) + 1:5000 rabbit anti-S235/236 phospho-S6 (Cell Signaling, 4857) + 1:2000 rabbit anti-TBC1D7 (Cell Signaling, 14949). The blots were incubated with primary antibody O/N at 4°C while shaking. The next day blots were washed 3 times in PBST and 1x in PBS followed by 1h incubation with 1:10,000 dilutions of secondary antibody IRDye 680RD Goat Anti-Rabbit IgG , Goat anti-Mouse IgG 800CW (Li-Cor biosciences). Blots were scanned with the Odyssey Clx (Li-Cor Biosciences).

#### Functional assay of ciliogenesis

Ciliogenesis was investigated in fibroblasts upon serum starvation as described previously (31). Briefly, fibroblasts were cultured in duplicate in DMEM at low FCS concentration (0.5 % (v/v)) on 24 mm cover slips in 12 wells plates for 48h. Next, cells were fixated with methanol and immunostaining was performed with primary antibodies: mouse anti-acetylated tubulin (Sigma Aldrich, T7451, ICC 1:8000) and rabbit anti-γ-tubulin (Sigma Aldrich, T3320, ICC 1:1000). Secondary antibodies used were: donkey anti-Mouse-Cy3 (ICC 1:200, Jackson Laboratories, 715-165-150) and donkey Anti-Rabbit DyLight 488 (ICC 1:3000) together with Hoechst (0.05 µg/ml) staining. Cilia lengths were counted blind and defined as normal (>3 μm) or short (<3 μm). A total of 100 cells were counted per cover slip. 50-80% of the skin-derived fibroblast should grow normal primary cilia under these conditions. Additional analysis of SHH signaling by purmorphamine mediated induction of *GLI* expression was performed as described previously (21). Briefly, fibroblasts were serum starved for 24h in DMEM (0.5 % (v/v) FCS). Thereafter, fibroblasts were treated with 20 μM purmorphamine (Sigma Aldrich®, SML0868) in DMEM (0.5 % (v/v) FCS) or DMSO control for 24h. RNA was isolated as above and reversed transcribed with TaqMan® Gene Expression Cells-to-CT™ kit (Thermo Fisher Scientific®). Relative *GLI1* expression was determined using TaqMan™ Gene Expression Assay (hGLI1 primer Hs01110766_m1 and housekeeping gene hCLK2 Hs00241874_m1) on a CFX96 Real-Time PCR Detection System (BioRad®). A fold change of between 4 and 40 after purmorphamine treatment for *GLI1* is considered normal.

## Results

### Diagnostic pipeline and user-friendly interface for RNA-seq expression outlier detection

We first compared global expression of genes known to be involved in NDD in diagnostic whole blood (collected in PAXgene blood RNA tube) and fibroblast RNA samples, and, consistent with previous work, found that on average approximately 10% fewer NDD exome panel genes were expressed in whole blood compared to fibroblast RNA (**Fig. S1A**) (12). As whole blood RNA-seq data also had more duplicates and smaller fragment size, indicating increased mRNA degradation, and lower transcript integrity number (TIN), we decided to use fibroblast RNA (**Fig. S1B-D**). To inhibit nonsense-mediated mRNA decay (NMD) and improve detection of abnormal transcripts, we also included RNA from fibroblasts cultured in the presence of cycloheximide (CHX). CHX treatment did not affect mRNA quality (**Fig. S1B-D**). We performed polyA-capture RNA-seq in 96 individuals including 67 with undiagnosed NDD, 10 with suspected neurofibromatosis type 1 (NF1), 16 individuals with undiagnosed non-neurological disorders and 3 healthy parents (**Table S2)**. In each case diagnostic WES analysis had failed to identify a causative variant (referred to as WES-negative) **(Fig. 1A)**. Outlier detection was performed by Z-score and p-value calculation for either genes (to detect abnormal expression), exons (to detect abnormal mRNA splicing including exon skipping) or introns (to identify intron retention and potential pseudo-exons) after OUTRIDER correction of RNA-seq counts (27). Data was made accessible via a self-developed user-friendly Jupyter-based web browser application (**Fig. 1C**). Candidate deviations in genes, exons or introns that were detected in the sample under investigation were visualized in volcano-, gene-fragment and exon ranking plots and selected for further analysis by direct inspection of mapped RNA-seq reads (e.g. BAM files) using the IGV (**Fig. 1C-6, 1C-7, 1C-8**). Finally, re-analysis of WES-data for rare variants, targeted Sanger sequencing, targeted testing of pre-mRNA splicing (RT-PCR, mini-gene exon trapping assay) and functional testing (e.g. Western blotting, immunostaining of cilia) were performed to help determine pathogenicity and provide a molecular diagnosis (**Fig. 1D**).

In the NDD cohort, a significant aberration in a known gene was identified in 9 NDD cases, for which further analysis showed a pathogenic or likely pathogenic variant, thereby providing a definitive diagnosis (**Table 1**). In 9/10 cases with suspected NF1, pathogenic variants in *NF1* were detected and have been summarized elsewhere (Douben et al. 2022; under revision). The *NF1* results were therefore not counted in the yield of the total NDD cases, but the RNA-seq data were used as controls.

**Table 1.**
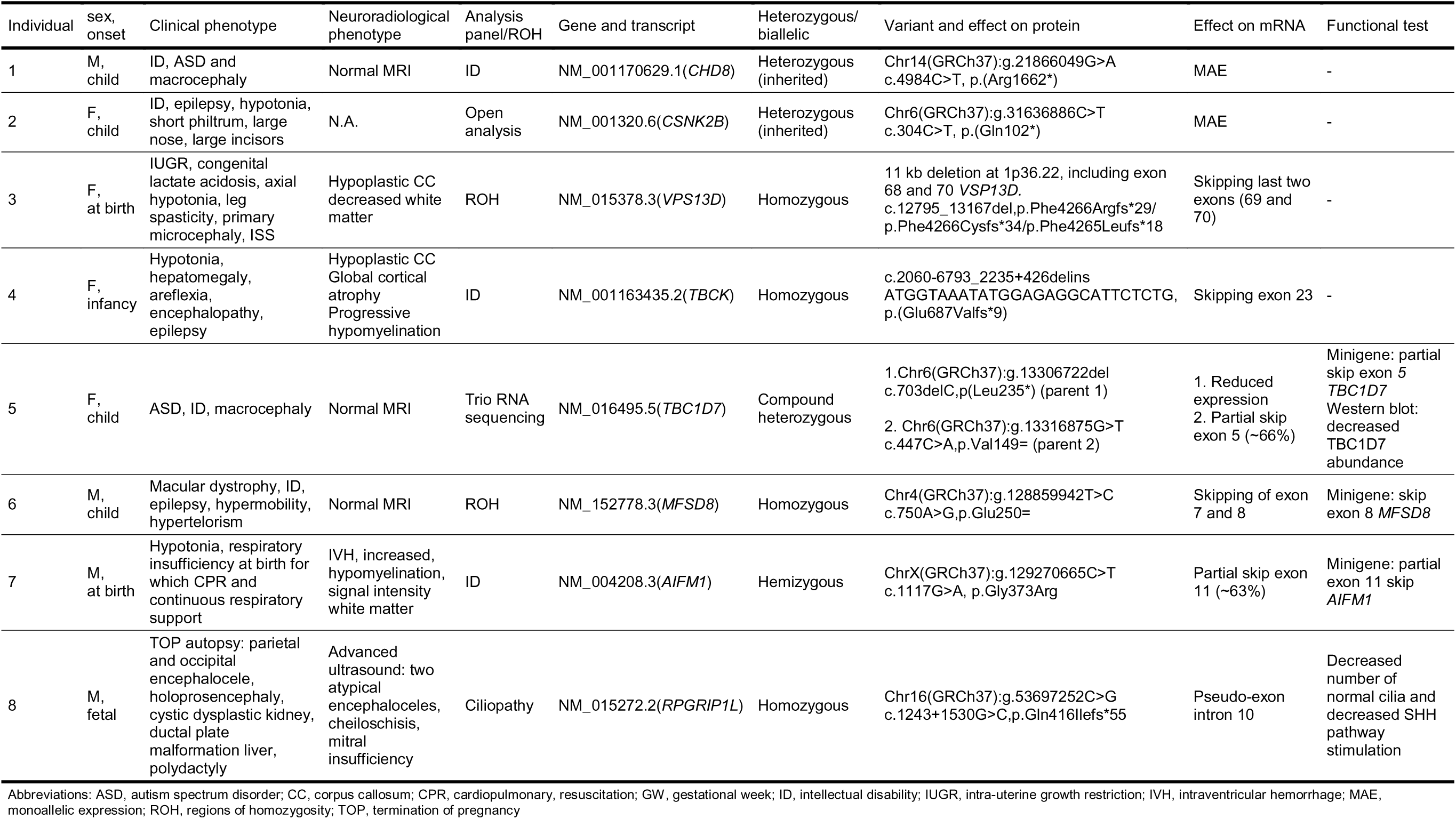
Overview of neurodevelopmental disorder cases where RNA-seq contributed to the identification of the causative variant

Phenotype descriptions are minimalized to avoid identification of the individual. More detailed phenotype descriptions are available on request.

### Detection of monoallelic expression of missed inherited variants in *CHD8* and *CSNK2B*

#### Case report 1

Individual 1 is a boy, born to non-consanguineous parents, and presented during childhood with an ID and autism spectrum disorder (ASD). Physical examination revealed dolichocephaly, large ears, slight asymmetry of hands (R>L) and feet (R>L), mild hypotonia and macrocephaly with an occipital frontal circumference (OFC) of 57 cm (+2.65 SD). He developed episodes of absences suspect for epileptic seizures, but with normal EEG. Brain MRI showed megalencephaly without structural abnormalities **(Fig. 2A,B)**. Analysis of *FRAXA* gene repeats was normal. WES trio analysis for *de novo*, homozygous, compound heterozygous and X-linked variants did not reveal a pathogenic variant. RNA-seq analysis at exon-level on ID panel genes revealed multiple downregulated exons of *CHD8* (**Fig. 2C,D**). Subsequent analysis showed allelic imbalance of *CHD8* due to a stop-gain variant NM_001170629.1(*CHD8*):c.4984C>T, p.(Arg1662*) **(Fig. 2E,F)**. Retrospective analysis of trio WES data revealed this stop-gain variant in both individual 1 and parent 2 (**Fig. 2F**). In RNA from untreated fibroblasts, 87% of the reads were from the wild-type allele and 13% contained the stop codon (**Fig. 2F**). In contrast, in RNA from fibroblasts treated with CHX the variant allele frequency (VAF) was ∼50%, indicating that the transcript containing the premature stop is subject to NMD (**Fig. 2F**). This *CHD8*:c.4984C>T, p.(Arg1662*) was previously reported as pathogenic (32, 33). Parent 2 of individual 1 is healthy, but macrocephalic (OFC 61.2 cm; +3.5 SD). Diagnostic DNA-methylation analysis using an lllumina lnfinium MethytationEplC Array showed a *CHD8-*related signature both in individual 1 and parent 2 (34). Carriers for pathogenic *CHD8* variants without a cognitive phenotype have previously been reported and therefore we conclude that the *CHD8* variant is causative for the NDD in individual 1 (35).

**Fig. 2.**
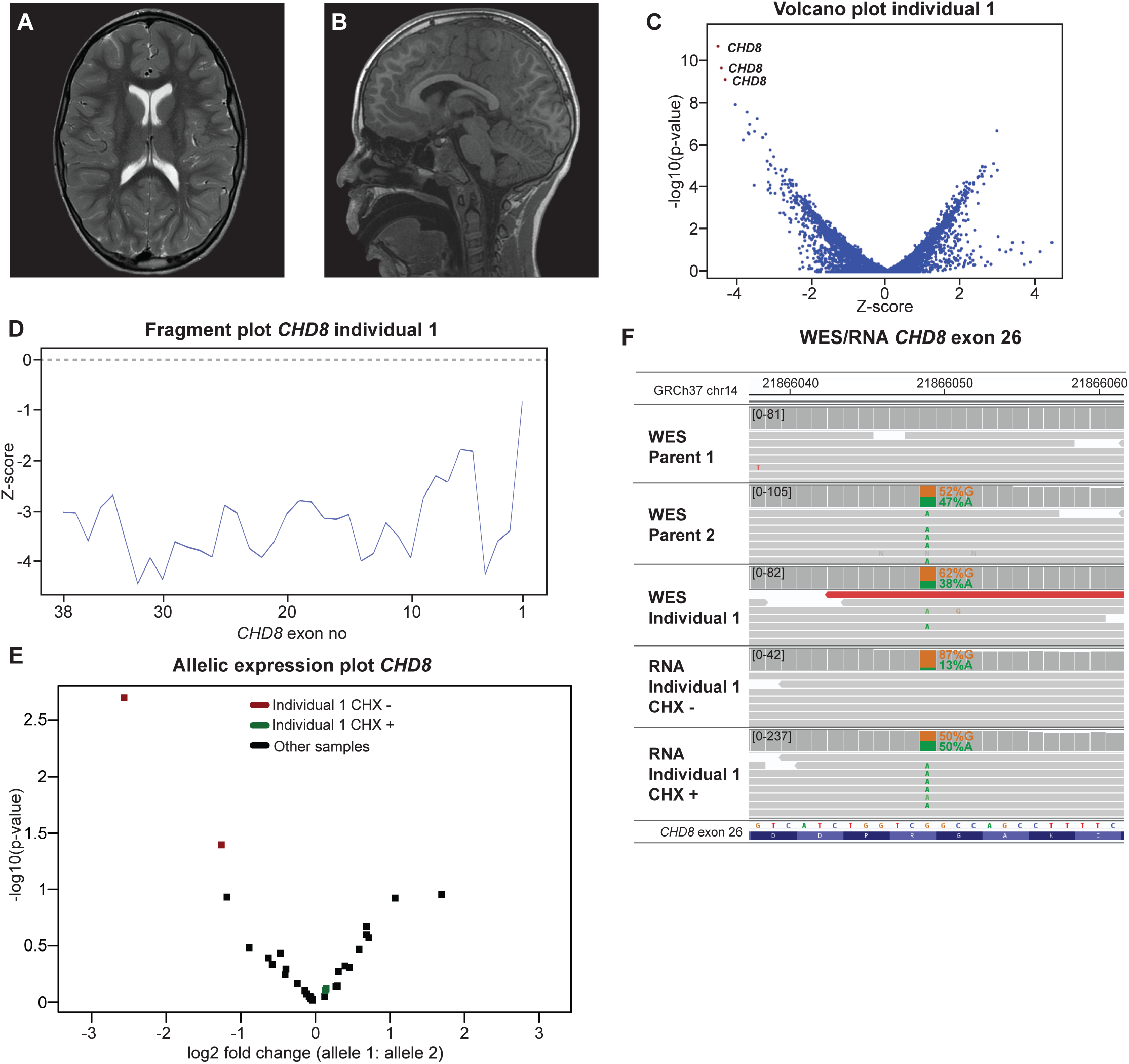
Monoallelic expression of *CHD8* due to an inherited stop-gain variant the cause of ASD and macrocephaly. **A, B** Individual 1’s brain MRI. Axial T2 **A** and sagittal T1 **B** weighted images showing megalencephaly with normal CSF spaces, without overt structural abnormalities. **C** Volcano plot at exon level (z-threshold=4; p-value<0.0025; ID panel) shows that multiple exons of *CHD8* are downregulated. **D** Individual 1’s fragment plot of *CHD8* of reveals that the majority of the exons have decreased expression. **E** Allelic expression plot of *CHD8*. Each dot shows the log2 fold change of one SNV calculated from the RNA-seq data reads. The two SNV with the highest allelic imbalance are from individual 1’s untreated fibroblast (red). **F** IGV plot of WES trio data (individual 1 and parents) and RNA-seq data (mapped reads) of untreated and CHX treated samples of individual 1 at the *CHD8* exon 26 region. MAE of *CHD8* is caused by an inherited stop-gain c.4984C>T,p.Arg1662*. RNA-seq from individual 1 shows monoallelic expression that is corrected by CHX treatment, confirming the sensitivity of the transcript containing the premature stop to NMD.

#### Case report 2

Individual 2, born to non-consanguineous parents, presented with epilepsy, moderate developmental delay, hypermobile joints and hypotonia. WES trio analysis, with the same variant filtering as used for case 1, did not reveal a causal variant. Full RNA-seq analysis at exon-level revealed decreased expression of all exons of *CSNK2B* (**Fig. S2A**). Subsequent analysis showed allelic imbalance due to an inherited stop-gain variant NM_001320.6(*CSNK2B*):c.304C>T, p.(Gln102*) (85% wild-type vs. 15% stop-gain variant in the untreated sample) (**Fig. S2B,C**). Heterozygous loss-of-function and missense variants in *CSNK2B* are associated with a comparable NDD (36–39). Individual 2 had a sibling with a similar, more severe, phenotype, who had died of pneumonia in adulthood. Re-analysis of WES-data confirmed that the heterozygous *CSNK2B*:c.304C>T, p.(Gln102*) was present in the sibling and heterozygous in DNA from blood and urine in parent 2. Neurological re-examination of parent 2, revealed behavioral problems, mild ID and premature memory loss, confirming the diagnosis in all three affected individuals.

### Detection of sub-gene deletions in *VPS13D* and *TBCK*

#### Case report 3

Individual 3 was born to healthy consanguineous parents after a pregnancy complicated by intra-uterine growth restriction, with a birth weight of 1490 g (p<3) and low Apgar scores (3-6 at 1 and 5 min). She presented with congenital microcephaly (OFC of 31 cm (-3.21 SD) at birth), lactate acidosis, type II atrial septal defect and failure to thrive. Brain MRI showed a hypoplastic corpus callosum and reduced white matter volume (**Fig. 3A,B**). Later she developed infantile spasms. Neurological examination showed axial hypotonia and spasticity of the legs. Full trio WES analysis and mitochondrial DNA sequencing did not reveal abnormalities. RNA-seq analysis focused on the exons of genes within regions of homozygosity (ROH), as identified by SNP-array, showed that the last two exons of NM_015378.3(*VPS13D*) were highly downregulated (exon 69 Z = -6.94; p = 1.07^-11^ and exon 70 Z = -6.74; p = 3.19^-10^) (**Fig. 3C-E**). Comparison of WES and RNA-seq data showed that no reads were mapped to exon 69 and 70 of *VPS13D*, indicating a homozygous deletion that was heterozygous in the parents. A genomic deletion within *VPS13D* Chr1(GRCh37):g.12559294_12573479delinsCAATGT was confirmed by PCR. (**Fig. 3F,G**). The effect of this genomic deletion is aberrant splicing from exon 68 to three different non-coding regions downstream of *VPS13D* (**Fig. S3)**. The result of these different splice events on protein level are three different frameshifts with a premature stop: p.(Phe4266Argfs*29), p.(Phe4266Cysfs*34) and p.(Phe4265Leufs*18). *VPS13D* encodes a protein implicated in supplying fatty acids to mitochondria, and bi-allelic variants in cause an NDD that matches the phenotype of individual 3 (40–44).

**Fig. 3.**
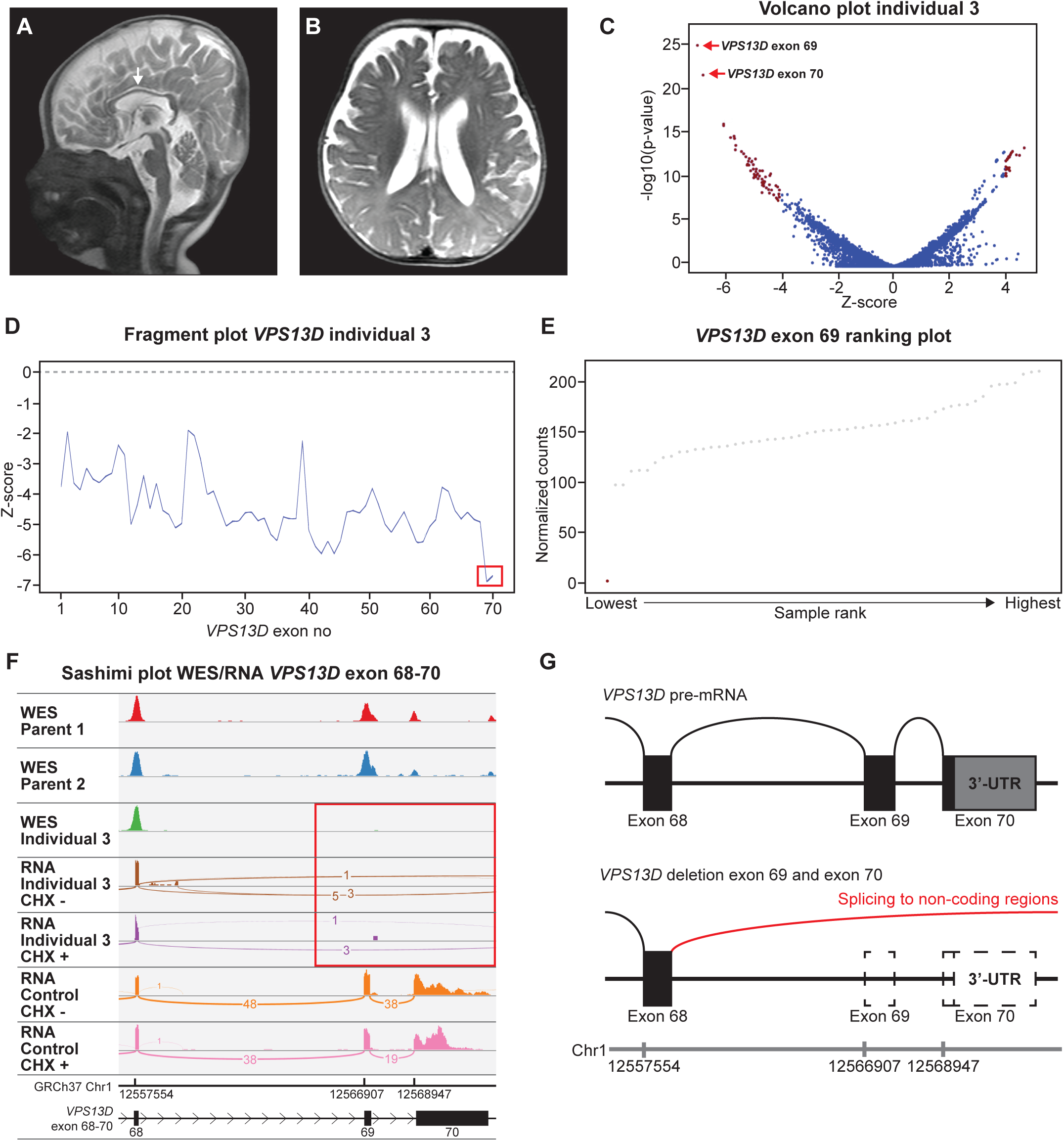
Deletion of the last two exons of *VPS13D* cause of an NDD. **A** Midsagittal T2-weighted image shows a hypoplastic corpus callosum (white arrow), and **B** Axial T2-weighted image shows moderate cerebral atrophy and decreased volume of the central white matter. **C** Volcano plot at exon level (Z-threshold=4; p-value<0.0025; no panel filtering).The two most downregulated exons are the last two exons (69 and 70) of *VPS13D* in the volcano plot. **D** individual 3’s fragment plot *VPS13D* shows a generalized decreased expression of *VPS13D*, with the last two exons having a Z-score of -6.94 (exon 69) and -6.74 (exon 70) (red box). **E** Exon ranking plot of exon 69 of *VPS13D.* Individual 3 (red dot) has close to zero read counts where other RNA samples (grey dots) have ≥ 97 counts. **F** IGV Sashimi plot of WES trio data (individual 3 and parents) and RNA-seq data (mapped reads) of untreated and CHX treated samples of individual 3 and a control at the *VPS13D* exon 68-70 region. Individual 3 has no DNA and RNA-seq reads mapped to *VPS13D* exon 69 and 70 (red box), due to a homozygous deletion of this exon. **G** Schematic representation of the effect of the genomic deletion in *VPS13D* in individual 3. Splicing can no longer occur from exon 68 to 69 and splicing towards non-coding regions is therefore seen from exon 68.

#### Case report 4

A girl, individual 4, born to non-consanguineous parents was hospitalized during infancy for increasing muscle weakness and head balance loss. Physical examination showed hypotonia, developmental delay and hepatomegaly. Disease progression caused areflexia and severe epileptic encephalopathy. Brain MRI revealed a hypoplastic corpus callosum, global atrophy and progressive hypo- and dys-myelination (**Fig. 4A**). WES trio analysis of ID panel and full exome was uninformative. RNA-seq analysis at exon-level on ID panel genes showed that exon 23 of NM_001163436.2(*TBCK*) was the most downregulated exon out of all transcriptomic exons detected (Z = -6.98; p = 5.22^-12^) (**Fig. 4B,C**), with close to zero reads compared to >100 reads in the rest of the cohort (**Fig. 4D**). Reanalysis of the WES data confirmed a lack of *TBCK* exon 23 coverage in individual 4, compatible with a homozygous genomic deletion, resulting in exon skipping and causing a frameshift and premature stop in the transcript: NM_001163436.2(*TBCK*):r.2060_2235del, p.(Glu687Valfs*9) (**Fig. 4E,F**). This deletion was confirmed by targeted PCR and sequencing: Chr4(GRCh37):g.107091826_107099220delinsATGGTAAATATGGAGAGGCATTCTCTG. Bi-allelic variants in *TBCK* cause a lysosomal storage-like disorder and a homozygous deletion of *TBCK* exon 23 has recently been described in two affected individuals, confirming the diagnosis in individual 4 (15, 45, 46).

**Fig. 4.**
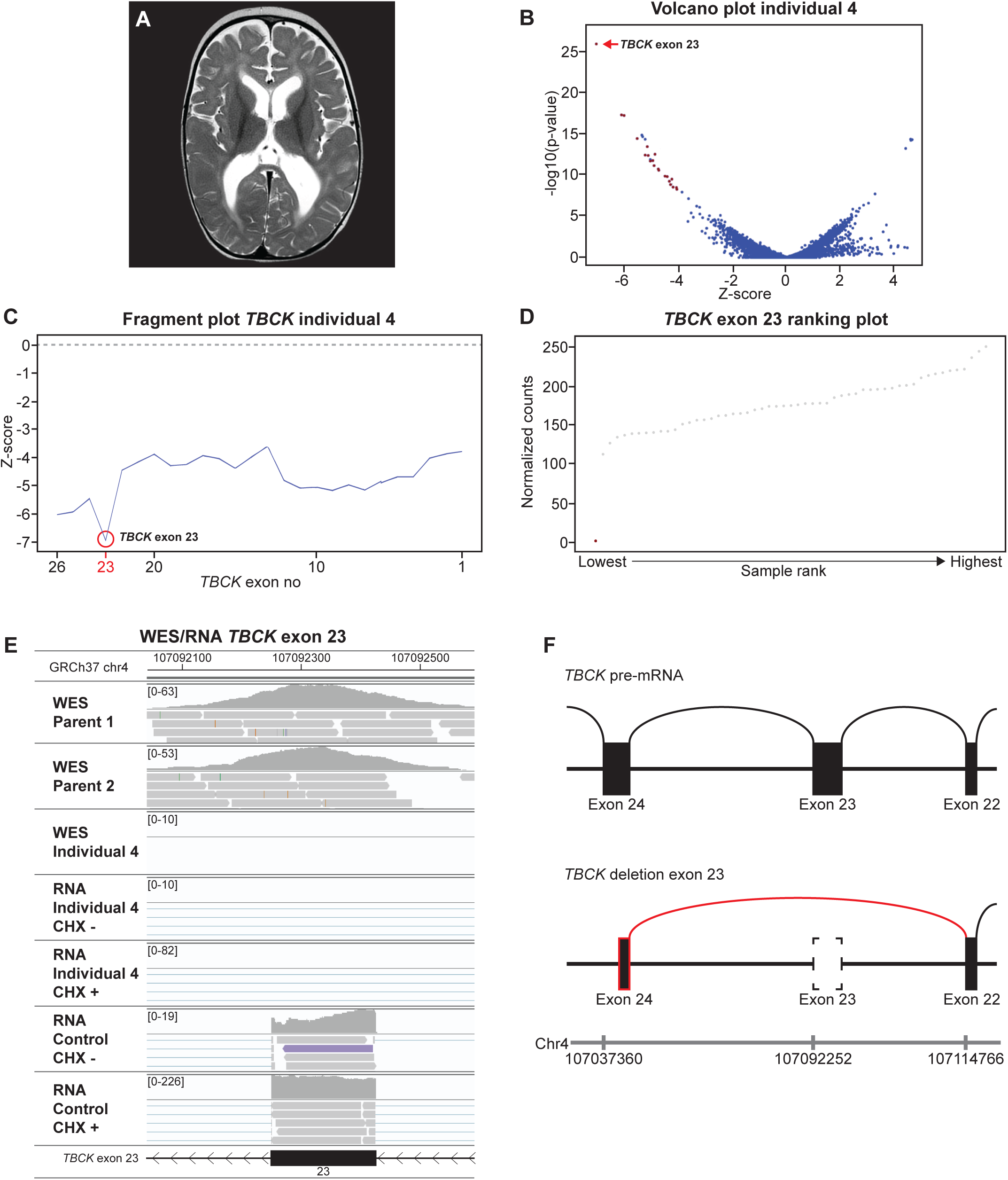
Homozygous deletion of *TBCK* exon 23 cause of an epileptic encephalopathy. **A** Axial T2 image showing cortical atrophy, enlarged lateral ventricles and decreased white matter. **B** Volcano plot at exon level (Z-threshold=4; p-value<0.0025; ID panel)). *TBCK* exon 23 is the most downregulated exon. **C** individual 4’s fragment plot of *TBCK* shows a complete downregulation of all exons of *TBCK*, where exon 23 is the most downregulated exon (Z-score=-6.98). **D** *TBCK* exon 23 ranking plot; individual 4’s RNA sample (red dot) has close to zero read counts, where all other RNA samples (grey dots) have ≥ 100 read counts. **E** IGV plot of WES trio data (individual 4 and parents) and RNA-seq data (mapped reads) of untreated and CHX treated samples of individual 4 and a control at the *TBCK* exon 23 region. Individual 4 has no DNA-seq and RNA-seq reads mapped to *TBCK* exon 23, due to a homozygous deletion of this exon. **F** Schematic representation of the outcome of the genomic deletion. Exon 22 splices to exon 24 resulting in a frameshift and truncated protein.

### Deleterious effect of synonymous variants on mRNA splicing (*TBC1D7* and *MFSD8*)

#### Case report 5

Individual 5, born to non-consanguineous parents, presented with unexplained macrocephaly (OFC: 57.1 cm; +2.6 SD), autistic traits and ID. Trio WES filtered for ID panel genes, *de novo,* X-linked and autosomal recessive diseases identified an inherited stop-gain variant NM_016495.5(*TBC1D7*):c.703delC, p(Leu235*). Biallelic variants in *TBC1D7* are associated with macrocephaly and ID (47, 48). Reanalysis of the WES data revealed a rare synonymous variant, *TBC1D7*:c.447C>A, p.(Val149=) (rs768742269) in exon 5 in individual 5 and parent 1. Trio RNA-seq analysis at exon-level showed downregulation of *TBC1D7* exon 5 in both parent 1 and individual 5 (**Fig. 5A and Fig. S4A**). The sequence reads revealed skewed allelic expression in parent 1 (VAF: 19%) and individual 5 (VAF: 37%) (**Fig. 5B**). Furthermore, both individual 5 and parent 1 showed skipping of exon 5, r.382_519del, p.(Glu128_Leu173del), hardly found in control samples (**Fig. 5C**). The synonymous *TBC1D7*:c.447C>A variant was predicted to disrupt an exonic splice enhancer (ESE) site **(Fig. S4B)** (49, 50). A mini-gene exon trapping assay confirmed that the variant causes skipping of exon 5 (**Fig. 5D,E).** The hypothesis that the two inherited *TBC1D7* variants were causative was further supported by decreased TBC1D7 protein abundance in fibroblasts derived from individual 5 (**Fig. 5F,G and Fig. S4C-I**). Furthermore, the TBC1D7 p.Glu128_Leu173del deletion prevents binding to the TSC complex (51). Nonetheless, consistent with the regulatory role of TBC1D7 in the TSC complex, we did not observe complete loss of inhibition of TORC1 signalling in fibroblasts from individual 5 (**Fig. S4H,I)** (52). Together, these data support the pathogenicity of the inherited synonymous *TBC1D7* variant.

**Fig. 5.**
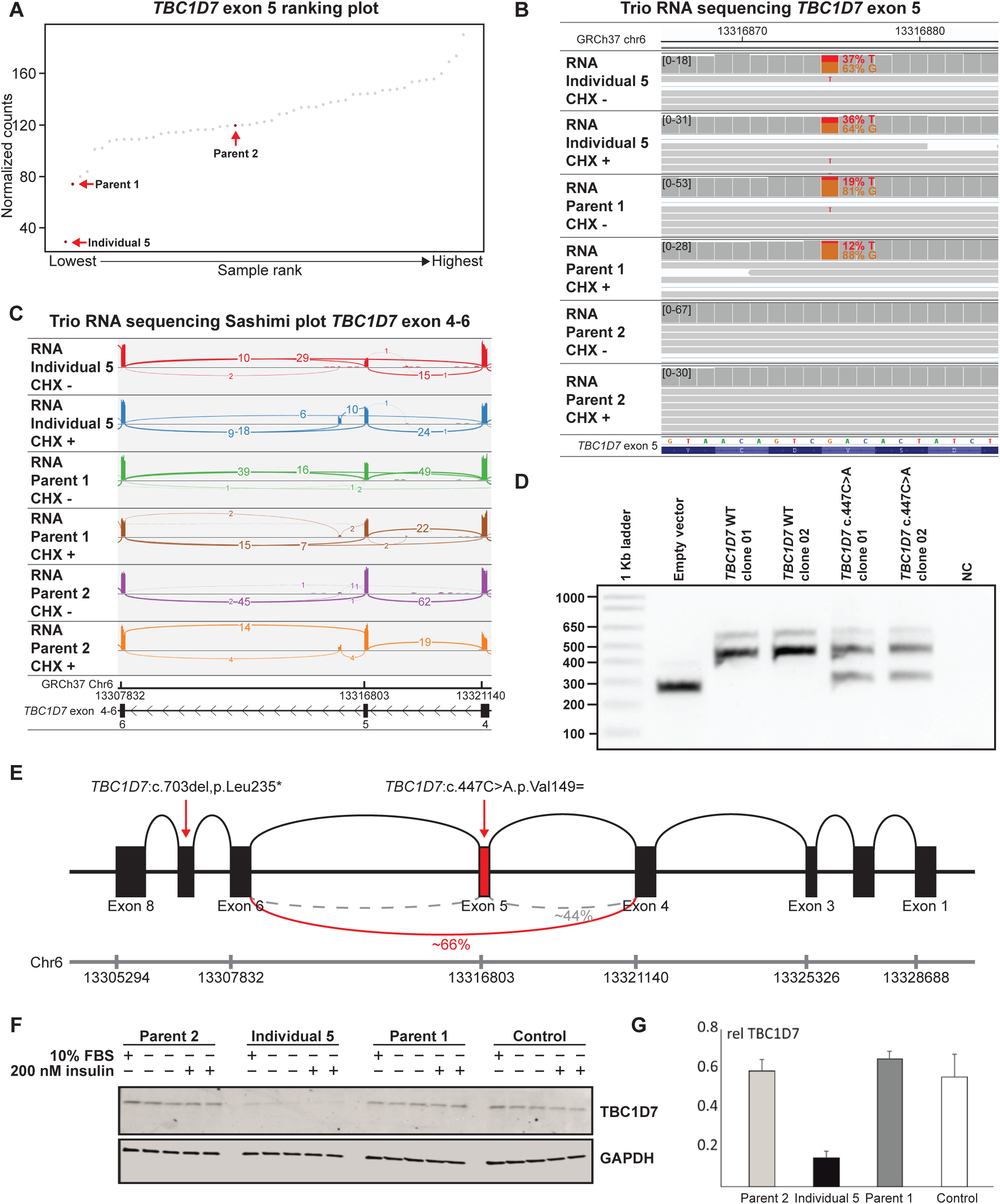
Trio RNA-seq reveals that a synonymous variant in *TBC1D7* has a deleterious effect on splicing. **A** *TBC1D7* exon 5 ranking plot: individual 5 and her parents. Individual 5 has a decreased expression of exon 5, where parent 1 has the second lowest expression. Parent 2 who is a carrier of a heterozygous premature stop (p.Leu235*) has normal expression levels of exon 5. **B** IGV plot of RNA-seq data (mapped reads) of untreated and CHX treated samples of individual 5 and her parents of *TBC1D7* exon 5 region shows allelic imbalance of the synonymous *TBC1D7*:c.447C>A,p.Val149= variant in parent 1, which is less pronounced in the individual 5. **C** Sashimi plot of RNA-seq data (mapped reads) of untreated and CHX treated samples of individual 5 and her parents of *TBC1D7* exon 4-6 region shows a partial skip of exon 5 in both parent 1 and individual 5, but not in parent 2. **D** Minigene exon trapping assay confirming that the synonymous variant TBC1D7:c.447C>A,p.Val149= induces a partial skip of exon 5 *TBC1D7.* **E** Schematic representation *TBC1D7* locus. Skipping of exon 5 is seen in 66% of the reads of individual 5. **F** Immunoblot analysis. Decreased TBC1D7 abundance is seen in the index, but not in parent 1 and 2. **G** Quantification of **F**, n=3.

#### Case report 6

Individual 6 is a boy, born to consanguineous parents, who had reduced visual acuity caused by macula dystrophy with subretinal debris in the macula (bull’s eye-like) with signs of progression. He was also diagnosed with mild ID and epilepsy during childhood. Physical examination revealed joint hypermobility and hypertelorism. Trio WES analysis of the ID panel and full exome analysis were uninformative. RNA-seq at exon-level focused on the genes within the ROH showed reduced expression of exons 7 and 8 of the transcript NM_152778.3(*MFSD8*) (**Fig. S5A-C**), due to skipping of either *MFSD8* exon 8 or exon 7 and 8 combined (**Fig. S5D)**. Skipping of exon 8 only, results in a frameshift, r.699_754del, p.(Arg233Serfs*5), whereas skipping of both exon 7 and 8 results in an in-frame deletion, r.554_754del, p.(Val185_Glu251del). Retrospective analysis of the WES data identified a homozygous synonymous variant NM_152778.3(*MFSD8*):c.750A>G, p.(Glu250=) in exon 8 that is predicted to affect splice enhancer sites (RESCUE-ESEs), leading to inactivation of the canonical donor site (spliceAI donor loss Δ-score:0.45; range: 0.00-1.00) (**Fig. S5E,F**) (50, 53). Indeed, mini-gene exon trapping analysis showed that the synonymous variant caused increased skipping of exon 8 (**Fig. S5G**). Pathogenic *MFSD8* variants cause an NDD or macular dystrophy (54, 55). Considering the phenotypic overlap, the variant was considered causative for the NDD phenotype of individual 6.

### Exonic splice enhancer variant in *AIFM1* causes exon skipping in X-linked progressive encephalopathy

#### Case report 7

Individual 7, was a male born with hypotonia, low Apgars (1-5-6 at 1, 5 and 10 minutes) and failed to breathe at birth. A mayo tube was inserted followed by cardiopulmonary resuscitation. Due to a persistent lack of a respiratory drive, intubation for mechanical ventilation was required. Brain MRI revealed a left-sided intraventricular hemorrhage, delayed myelination and increased white matter signal intensity more pronounced in the left temporal-occipital lobe **(Fig. 7A,B).** Due to the combination of symptoms a metabolic or mitochondrial disease was suspected. Blood lactate was elevated, and increased excretion of lactate and pyruvate in urine was detected together with increased levels of 2-hydroxybutyric acid, 3-hydroxyisobutyric acid, fumaric acid and malic acid. SNP-array and trio WES analysis of ID panel filtered for *de novo,* recessive and X-linked variants did not identify a cause. RNA-seq analysis at gene-level filtered for ID panel genes showed reduced expression of NM_004208.3*(AIFM1)* (chromosome Xq26.1) **(Fig. 6C)**. Reduced expression was explained by partial skipping of exon 11, found in 63% of reads in the CHX treated sample, resulting in a frameshift and premature stop (r.1076_1164del,p.(Glu359Glyfs*4)) **(Fig. 6D)**. Exon 11 contained a an inherited nonsynonymous variant c.1117G>A,p.(Gly373Arg) not present in GnomAD v3.1, however the amino acid change was predicted to have no impact on the protein by Alamut software missense prediction tools **(Fig. 6E)**. However, this variant is predicted to result in an ESE loss (RESCUE-ESE), leading to a loss of the canonical exon splice sites (spliceAI acceptor loss: Δ-score:0.37; spliceAI donor loss Δ-score:0.40; range: 0.00-1.00), which was confirmed using a minigene exon trapping assay **(Fig. 6F-H).** AIFM1 facilitates the assembly of the mitochondrial complex I and plays an important role in caspase-independent apoptosis (56, 57). Loss-of-function *AIFM1* variants are associated with variable disease, including progressive encephalopathies. The phenotype in individual 7 matches the most severe cases described in literature (58, 59).

**Fig. 6.**
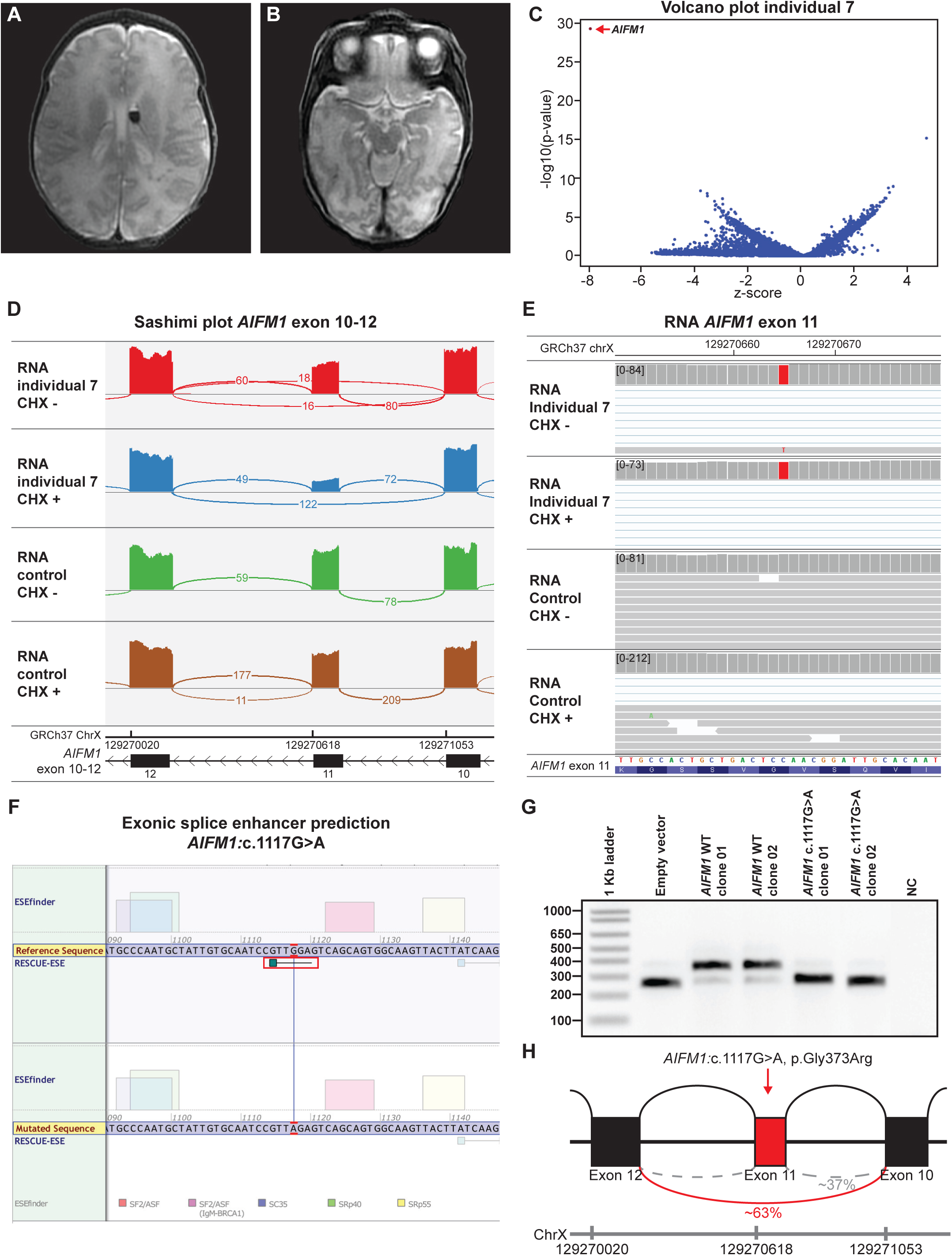
*AIFM1* non-synonymous variant causes partial exon skipping in an X-linked disease. **A, B** Individual 7’s brain MRI scan. Axial T2 weighted images showing delayed myelination and hemosiderine signal in the left ventricle, increased signal intensity of the white matter more pronounced in the posterior left temporal-occipital area, compared to the right lobe. **C** Volcano plot at gene level (Z-threshold=4; p-value<0.0025; ID panel). The only downregulated gene is *AIFM1.* **D** Sashimi plot of RNA-seq data (mapped reads) of untreated and CHX treated samples of individual 7 and a control of *AIFM1* exon 10-12 showing a partial skip of exon 11 in individual 7. **E** IGV view of RNA-seq data (mapped reads) of untreated and CHX treated samples of individual 7 and a control of *AIFM1* exon 11 showing a homozygous missense variant c.1117G>A,p.Gly373Arg. **F** Alamut splice enhancer prediction for the c.1117G>A,p.Gly373Arg shows loss of a RESCUE-ESE. **G** Minigene exon trapping assay confirms that the *AIFM1* missense variant c.1117G>A,p.Gly373Arg increases the likelihood of exon skipping. **H** Schematic representation of the effect of the *AIFM1* missense variant on splicing. 63% of the reads show (based on CHX treated cells) skipping of exon 11 in individual 7.

**Fig. 7.**
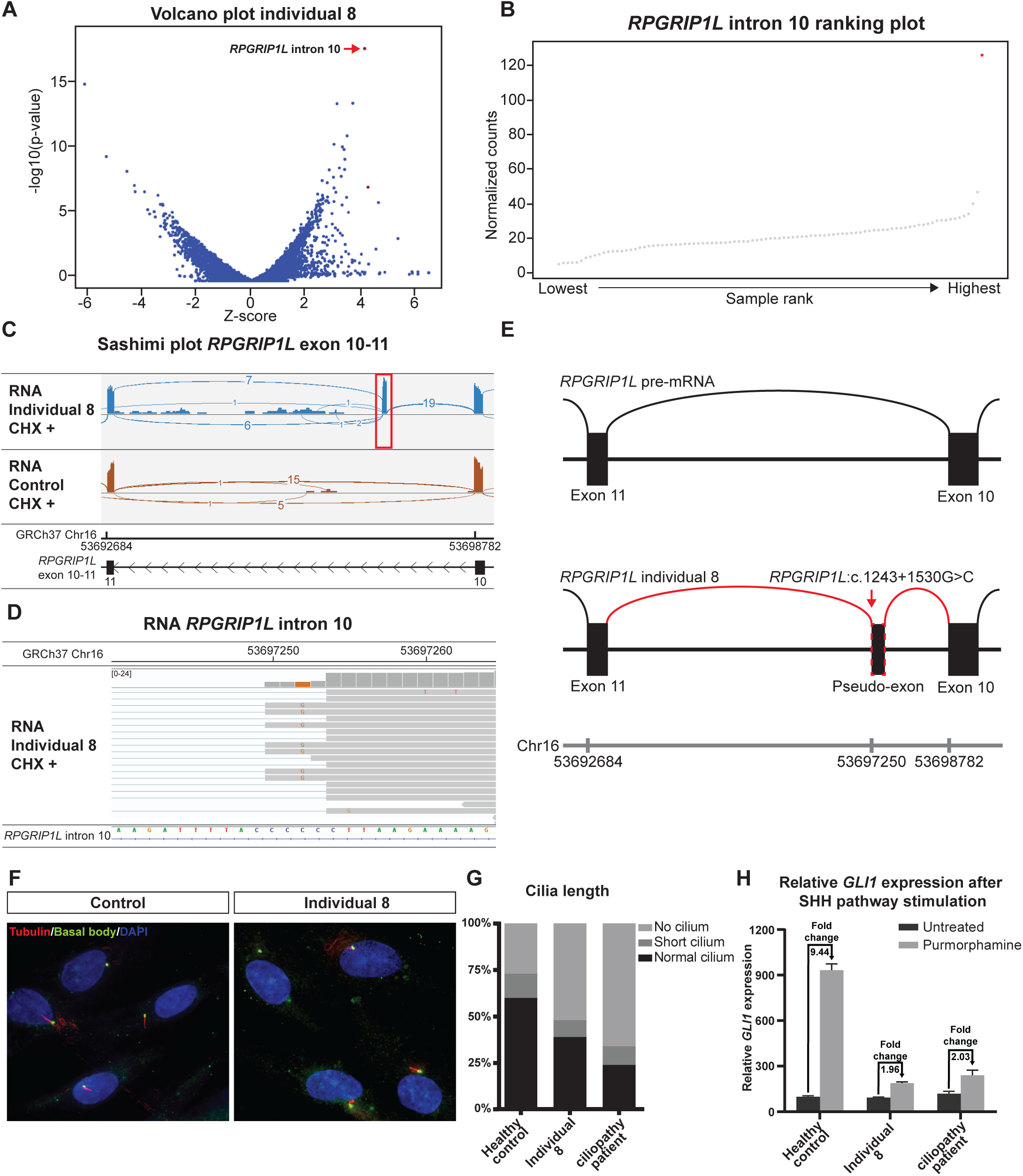
Non-coding variant in *RPGRIP1L* results in a pseudo-exon inclusion. **A** Volcano plot at intron level (Z-threshold=4; p-value<0.0025; ciliopathy panel) showing that the most upregulated intronic region maps to *RPGRIP1L* intron 10. **B** *RPGRIP1L* intron 10 ranking plot; individual 8’s RNA seq sample (red dot) has >120 reads mapped to this region where other samples (grey dots) have on average ∼20 normalized counts. **C** Sashimi plot RNA-seq data (mapped reads) of CHX treated samples of individual 8 and a control of *RPGRIP1L* exon 10-11 shows complete splicing towards pseudo-exon from exon 10 in individual 8 (red box). **D** IGV view of individual 8’s RNA-seq sample (CHX treated) zoomed in to the *RPGRIP1L* pseudo-exon that contains a homozygous c.1243+1530G>C variant in close proximity to the splice donor site. **E** Schematic representation of the effect of the intronic c.1243+1530G>C variant in *RPGRIP1L*. A pseudo-exon is included in the *RPGRIP1L* transcript between exon 10 and 11. **F** Immunostaining of acetylated tubulin and γ-tubulin (basal bodies) showing a decreased number of cells with normal cilia in individual 8. **G** Quantification of **F**, 50-80% of cells should grow a normal primary cilium. **H** RT-qPCR of *GLI1* after SHH pathway stimulation with purmorphamine. Individual 8 has a similar fold change *GLI1* expression as a known ciliopathy patient (1.96x and 2.03x)

### Non-coding variant causing inclusion of a novel pseudo-exon in *RPGRIP1L*

#### Case report 8

In a pregnancy of a consanguineous couple, advanced ultrasound revealed a fetus, individual 8, with an asymmetrical parietal and occipital encephalocele, polydactyly and hydronephrosis. Due to the severe fetal malformations, the parents decided to terminate the pregnancy. Pathological examination confirmed the ultrasound abnormalities. Trio WES and SNP array analysis did not identify a pathogenic variant. RNA-seq analysis at intron-level focusing on ciliopathy panel genes revealed a highly expressed intronic region mapping to intron 10 of NM_015272.2(*RPGRIP1L*) **(Fig. 7A,B)**. The expressed intronic region was caused by the inclusion of a pseudo-exon between exon 10 and 11 of the *RPGRIP1L* transcript, r.1243_1244ins1243+1459_1243+1532, p.(Gln416Ilefs*55), most clearly visible in RNA-seq data of CHX treated fibroblasts **(Fig. 7C)**. Incorporation of the pseudo-exon in the *RPGRIP1L* transcript was due to a homozygous c.1243+1530G>C variant, that is predicted to create a non-canonical splice donor site (spliceAI donor gain Δ-score:0.64) **(Fig. 7D)**. Biallelic, loss-of-function *RPGRIP1L* variants (OMIM:610937) have been associated with Meckel syndrome 5 (OMIM:611561) (60). We did not observe *RPGRIP1L* transcripts lacking the pseudo-exon in individual 8, indicating that the c.1243+1530G>C variant results in a complete loss of RPGRIP1L function **(Fig.7E)**. We assessed the morphology of cilia in fetal fibroblasts from individual 8 after serum starvation. We observed a decreased number of normal cilia (control=60% vs. *RPGRIP1L* index=39%; normal range = 50-80%) and an increased number of cells lacking a cilium (control=28% vs *RPGRIP1L* index=52%) (**Fig. 7F,G**). Furthermore, treatment with the Sonic Hedge Hog (SHH) agonist purmorphamine, which induces cilium-dependent induction of *GLI1* expression, failed to induce *GLI1* expression in fibroblasts from individual 8 (**Fig. 7H**). The RNA-seq results and ciliary defects confirm that the deep-intronic *RPGRIP1L* variant in individual 8 is pathogenic.

### Pathogenic splicing in *DCC*; a gene not expressed in fibroblasts

#### Case report 9

Individual 9, born to consanguineous parents, presented with a global developmental delay, unsteady gate, pyramidal signs, oculomotor apraxia, speech delay and apraxia of the hands. Brain MRI showed agenesis of the corpus callosum, elongated mesencephalon and a thin pons with a midline posterior cleft, indicating a decussation defect of pontine cross fibers. MRI indicated a multiple commissural defect, typically described for individuals with biallelic loss-of-function variants in *DCC* **(Fig. 8A,B)** (61). *DCC* was present in an ROH and was fully sequenced at exon level, but no pathogenic variants were identified. WES panels for MCD, ID, and trio full exome analysis were also uninformative. RNA-seq analysis was performed but expression of *DCC* was not detectable in either cultured fibroblasts or blood (**Table S3**). RNA-seq identified decreased expression of NM_003184.3(*TAF2*) (**Fig. 8C**). In individual 9’s cells treated with CHX a pseudo-exon between *TAF2* exon 19 and 20, r.2558_2559ins2558+643_2558+808, was observed in 35% of the reads **(Fig. 8D)**. RNA-seq also revealed a homozygous variant in the pseudo-exon, corresponding to *TAF2* (Chr8(GRCh37):g.120773964C>G,c.2558+691G>C), which was predicted to activate the non-canonical splice acceptor site at c.2558+643 (spliceAI acceptor gain Δ-score:0.22; range: 0.00-1.00), resulting in a frame shift and premature stop (p.*(Ser853Argfs*28)) **(Fig. 8E)**. The *TAF2* variant mapped to an ROH and was not reported in GnomAD v3.1. Pathogenic *TAF2* variants cause moderate/severe NDD and a thin or absent corpus callosum (OMIM: 614340). The *TAF2* variant was a possible candidate although it was not fully explanatory for the full agenesis of the corpus callosum and the pons abnormality of individual 9 (62, 63). Furthermore, we still observed normal splicing in 65% of the reads and we were puzzled by the fact that *TAF2* tolerates heterozygous loss-of-function variants (pLI = 0; gnomAD v.2.1.1). Therefore, we suspected that the reduction in functional TAF2 due to abnormal *TAF2* mRNA transcript may not be sufficient to be pathogenic Therefore, we conducted RT-PCR, allowing detection of low expression levels not detected by RNA-seq using our parameters, of *DCC* on cDNA obtained from fibroblasts of individual 9. Surprisingly, we found an increased amplicon size of *DCC* exon 25 and 26, and by Sanger sequencing we confirmed the presence of a pseudo-exon (r.3736_3737ins3736+8518_3736+8576) **(Fig. 8F,G)**. Subsequent genomic DNA Sanger sequencing identified a homozygous deep intronic variant in intron 25 of NM_005215.3(*DCC*): Chr18(GRCh37):g.51002897T>G, c.3736+8517T>G, resulting in the activation of a strong splice acceptor site **(Fig. 8H)**. The inclusion of this pseudo-exon results in a frameshift (p.(Ala1246Glyfs*23)) therefore a truncated DCC protein, which fully explains individual 9’s phenotype (61).

**Fig. 8.**
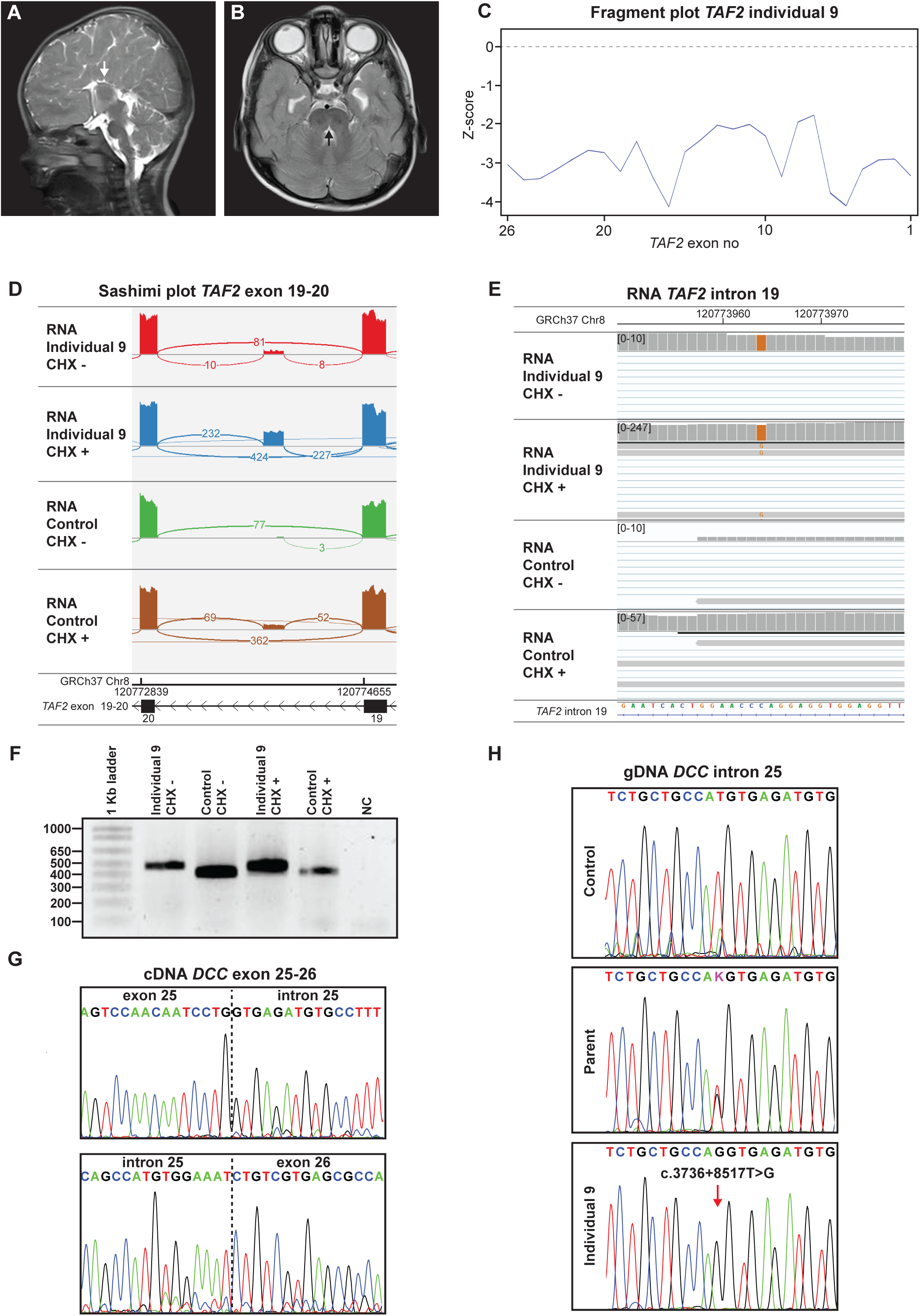
Benign pseudo-exon in candidate gene *TAF2*, in an individual with pathogenic splicing in *DCC,* that is not expressed in fibroblasts. **A, B** Individual 9’s brain MRI. **A** Sagittal T2 weighted image at midline shows an elongated mesencephalon and a complete agenesis of corpus callosum (white arrow). **B** Axial T2 images showing a thin pons with a posterior midline cleft (black arrow). **c** Individual 9’s fragment plot of *TAF2* showing that the whole gene is downregulated. **D** Sashimi plot of RNA-seq data (mapped reads) of untreated and CHX treated samples of individual 9 and a control of the region *TAF2* exon 19-20 shows that inclusion of the pseudo-exon between exon 19 and 20 of *TAF2* occurs in approximately 35% of the reads (CHX treated fibroblast) in individual 9 and at a much lower frequency in control. **E** IGV view of RNA-seq data (mapped reads) of untreated and CHX treated samples of individual 9 and a control zoomed in on the region of the pseudo-exon located in intron 19 of *TAF2*. A homozygous NM_003184.3(*TAF2*):c.2558+691G>C is present in this pseudo-exon. **F** Agarose gel result of RT-PCR product of exon 25-26 *DCC* of untreated and CHX treated fibroblasts of individual 9 and a control. RT-PCR shows an increased product size in individual 9’s products. **G** Sanger sequencing results of **F** individual 9’s RT-PCR (untreated) sample reveals inclusion of a pseudo-exon between exon 25 and 26 in NM_005215.3(*DCC*). **H** Genomic DNA Sanger sequencing of the *DCC* intron 25 region in gDNA of a control individual, individual 9’s parent and individual 9 reveal the presence of a deep intronic homozygous c.3736+8517T>G variant in individual 9, absent in the control gDNA and heterozygous in the parent. This results in the activation of a splice acceptor site.

## Discussion

Here, we show that implementation of RNA-seq analysis complements the routine genome diagnostic work-up of molecularly undiagnosed NDD individuals. In our cohort, we performed RNA-seq on cultured fibroblasts derived from skin biopsies, untreated or treated with cycloheximide (CHX), followed by a OUTRIDER Z-score-based outlier analysis of expressed transcripts, exons and introns, integrated into a user-friendly web-based application to facilitate data analysis. In our cohort of 67 NDD individuals we reached a definitive diagnosis in 8 cases using RNA-seq analysis, with a total yield of 12%, which is in the same range as previously published studies on RNA-seq for other groups of Mendelian disorders (8, 9, 11–16).

The cases presented here highlight that a wide range of genetic mechanisms, in a group of diverse NDDs, can be elucidated by RNA-seq. Our approach not only shows the complementary contribution of RNA-seq analysis to DNA diagnostics, but also the added value of integrating reverse neurological clinical phenotyping in the diagnostic workup of undiagnosed NDD in a multidisciplinary team. The pathogenic abnormalities identified in this NDD cohort included MAE of missed inherited variants (case 1 and 2), small genomic deletions (case 3 and 4), non-coding variants causing transcriptional inclusion of a pseudo-exon, also known as a poison exon (case 8 and 9), or (non)-synonymous variants, originally predicted to be benign, but by RNA-seq shown to result in skipping of the exon containing the variant (case 5, 6 and 7).

We found that, when using fibroblasts as a source of RNA instead of whole blood RNA, RNA and data quality was higher, based on our control parameters, and more genes known to be involved in NDD could be detected. This matches a recent study showing that fibroblasts have broader gene expression than whole blood, potentially allowing identification of mis-splicing events in more individuals (64).

Furthermore, individuals described here had pathogenic variants in genes with good expression in fibroblasts, but almost all showing low or variable expression in blood including *VPS13D*, *TBCK*, *MFSD8*, *AIFM1* and *RPGRIP1L*.

We used fibroblasts cultured in the presence and absence of CHX and found that both conditions provided insight critical to variant identification and classification. Untreated fibroblasts allowed identification of a drop in expression due to NMD, as seen in case 6, 7 and 9, whereas the abnormal transcript could be readily identified in the data from the CHX treated cells. This helped to determine the frequency of normal vs. abnormal splicing events, when for example the exon skip is only partial (case 7 and 9). CHX treatment can also help identify allelic imbalance, as seen in cases 1 and 2. Thus, including CHX treated samples provides useful additional insights when applying RNA-seq in genome diagnostics. We adopted exon- and intron-level Z-score-based outlier analysis, in addition to gene outlier analysis, allowing us to detect abnormal mRNA splicing (exon skipping/pseudo-exons) in the absence of changes on the expression levels of a gene. This increased the resolution and immediately helped with the identification of the relevant locus in raw RNA/DNA-seq data, the effects on transcription and the causative rare variant. Our diagnostic RNA-seq pipeline serves as a complement to the recently published DROP pipeline, which uses OUTRIDER for outlier expression at gene level, and FRASER, an algorithm based on the detection of splicing events based on split reads for the detection of aberrant splicing (28, 65). We have also tested FRASER on our RNA-seq data and although both FRASER and our exon- and intron-level Z-score-based approach detected the aberrant splicing events, FRASER resulted in detection of more background splice changes and/or false positives. This is partly because in CHX treated samples, abnormal splicing events that occur under physiological conditions—and are normally subject to NMD—are detected well. Furthermore, we focused on making the analysis interface user-friendly and easily accessible, with tunable settings for filtering, including selection of ROH, specific gene panels, Z-score/p-value cutoffs and the output of gene lists. Our method also offers a range of graphical visualizations assisting in determining data quality (e.g. PCA plot) and identifying the nature of outlier expression (e.g. Volcano plots, exon ranking plots, chromosomal Z-score plots).

Our study highlights several findings relevant to those investigating the genetic causes of NDD, showing multiple types of pathogenic mechanisms that we detect by RNA-seq in undiagnosed individuals. We report small DNA deletions in known disease genes, which were missed by routine WES (case 3 and 4). Some deletions could have been detected by CNV analysis of WES data—provided that there is sufficient coverage—, but this was not available at the time WES analysis was performed for these individuals. Nonetheless, even if these variants had been identified by WES analysis, they still required additional analyses to establish pathogenicity, and RNA-seq provided a complementary assay in these cases. We also describe dominant pathogenic truncating variants (case 1 and 2), which were excluded during WES trio filtering, and were instead detected by RNA-seq as MAE and reduced expression of the gene. Both variants were inherited from an apparently healthy parent and were therefore excluded during WES trio filtering. Trio analysis of the complete exome or of large gene panels has been extremely fruitful in diagnosis of ID, where *de novo* (often private) variants account for up to 30% of the causes (66). Missing inherited, putatively pathogenic variants can be overcome by the use of algorithms, e.g. based on HPO terms, for variant selection.

In case 8, where there was a suspicion for a ciliopathy, we identified a pseudo-exon explained by a deep intronic, homozygous variant. In principle, such pathogenic variants would be detected by WGS, but the interpretation of their effect can often not be predicted effectively, and therefore it is necessary to have a complementary functional RNA readout such as RNA-seq. Similarly, the added value of RNA-seq is especially evident in cases of (non)-synonymous variants that influence splicing independent on canonical splice donor/acceptor sites (case 5, 6 and 7). The homozygous *MFSD8* synonymous variant (individual 6) is an illustrative example of a variant with severe clinical consequences, which would not have been detected with available diagnostic techniques. Such examples stress the utility of RNA-seq analysis for the interpretation and prioritization of rare variants on splicing that would otherwise be classified as (likely) benign based on DNA-seq analysis alone. For these three cases 5,6, and 7 the pathogenicity of the variant could be explained by ESE(s) loss. However, interpretation of such variants can be difficult, as seen in case 6, where the ESE prediction tools (RESCUE-ESE, ESEfinder) provided conflicting predictions (49, 50). Although the recently developed spliceAI tool shows potential in predicting the effect on splicing for coding variants (case 6, 7) and non-coding variants (case 8, 9), it did not predict skipping of *TBC1D7* exon 5 (case 5) (53). Thus, we highlight four types of pathogenic variants that are effectively detected by RNA-seq in NDD: small deletions, MAE, pseudo-exons caused by deep-intronic variants and hard-to-prioritize/interpret variants affecting splice enhancers. Therefore, RNA-seq is a very useful complementary technique to identify pathogenic variants in undiagnosed NDD in routine DNA diagnostics.

Although RNA-seq improves the diagnostic yield, there are also pitfalls related to the identification of novel variants of unknown significance and to limited expression of candidate disease genes. Even though up to ∼83% of genes in, for example, the ID panel have sufficient expression in fibroblasts, ∼17% of genes relevant to NDD—including *DCC* as described in case 9—are not expressed in fibroblasts (**Table S3**). In the future, the use of a mRNA capture for lowly expressed genes in fibroblasts or patient-derived trans-differentiated/iPSC-derived neurons may overcome this pitfall and would allow detection of abnormally expressed/mis-spliced genes that have no expression in clinically accessible tissues (67).

We emphasize that even after analysis of both DNA and RNA, complementary functional tests are sometimes required to confirm pathogenicity. We find that mini-gene exon trapping assays is often necessary to confirm that a specific variant of interest causes aberrant splicing (case 5, 6 and 7). For case 8, suspected for a ciliopathy, we were able to directly test the cilia morphology and function to complement the clinical diagnosis. Reverse phenotyping—i.e. multidisciplinary team re-evaluation of an individual phenotype after multi-omics tests—becomes increasingly relevant in the modern genome-first diagnostic approach to NDD and was instrumental for the identification of the deep-intronic pathogenic *DCC* variant in our cohort (68). Altogether, accurate clinical/reverse phenotyping, DNA- and RNA-based diagnostics and functional testing complement each other and are all essential in providing a definitive diagnosis in an individual with a rare genetic disorder (69).

As our study and others show, RNA-seq increases the diagnostic yield typically by ∼10-20%. To obtain a definitive diagnosis in all individuals with a genetic disorder, additional tests are required, and WGS alone appears not be sufficient. For example, proteome analysis can increase the diagnostic yield where both DNA and RNA analysis fail to identify a pathogenic change (70). Introduction of specific epigenetic methylation signatures for NDD, as a complementary diagnostic aid, could also be valuable (71). It is clear that there currently is no single technology capable of serving all diagnostic queries and solving all genetically undiagnosed NDD cases. As proposed for cortical malformations, a multilevel assessment of disease process, taking into account clinical, genome-diagnostic and functional data, is needed to identify disease cause and pathogenesis (72).

## Conclusions

RNA-seq and complementary functional tests are an essential component in the diagnostic workflow of individuals with an NDD where DNA-diagnostics failed to establish a molecular diagnosis. As the number of (non-)coding variants will only increase with the wider implementation of WGS, RNA-seq will continue to aid in the identification and interpretation of unexpected pathogenic variants affecting mRNA processing in genetic disorders.

## Data Availability

All data produced in the present study are available upon reasonable request to the authors (IRB protocol METC-2012-387)

## Abbreviations

ASD: Autism spectrum disorder
CHX: Cycloheximide
ESE: Exonic splice enhancer
ID: Intellectual disability
MAE: Monoallelic expression
NDD: Neurodevelopmental disorder
NGS: Next generation sequencing
NMD: Nonsense-mediated mRNA decay
OFC: Occipital frontal circumference
RIN: RNA integrity number
RNA-seq: RNA sequencing
ROH: Regions of homozygosity
SNP: Single nucleotide polymorphism
SNV: Single nucleotide variant
TIN: Transcript integrity number
VUS: Variant of uncertain significance
WES: Whole exome sequencing
WGS: Whole genome sequencing

## Supplementary information

**Suppl. Tables: Table S1.** List of genes NDD panels Erasmus MC. **Table S2.** Overview samples RNA seq cohort. **Table S3.** Expressed and not expressed genes in fibroblasts or blood for NDD gene panels. **Suppl. Figures: Fig.S1** Gene panel expression and quality control of fibroblast and PAX. **Fig. S2.** Mono-allelic expression of *CSNK2B*. **Fig. S3.** Splicing from VPS13D exon 68 occurs to three different non-coding regions in individual 3. **Fig. S4.** TBC1D7 western blot and TSC-mTOR pathway analysis**. Fig. S5.** Synonymous variant *MFSD8* causes exon skipping.

## Declarations

### Ethics approval and consent to participate

For all included individuals informed consent for all diagnostic tests and publication was obtained (IRB protocol METC-2012-387).

### Consent for publication

All individuals or legal guardians provided written consent to share anonymized clinical and analysis data.

### Availability of data and materials

All WES and RNA-seq data are stored in a repository at the Clinical Genetics in het Erasmus MC. The consent agreement only allows us to share anonymized patient data, therefore we are unable to share the BAM or VCF files. Count data and our Jupyter-based web browser application for RNA-seq analysis is available on request/will be made available online.

### Competing interests

The authors declare to have not competing interest.

### Funding

TJvH was funded by an Erasmus University Rotterdam (EUR) fellowship.

### Authors’ contributions

Conceptualization: JD, RS, GMSM, TJvH. Methodology: JD, RS, MB, WGdV, MMvVP, KM, HD, PE, EK, LMAvU, GG, JJJ, FWV, MLTvdS, FSN, MW, LHH, MHW, MN, TJvH. Formal analysis and investigation: JD, RS, MB, WGdV MMVP, KM, HD, PE, EK, LMAvU, GG, YvI, FWV, MLTvsS, FSN, HHH, MW, VJMV, MJ, AJAK, IMBHL, MHW, MN, GMSM. Writing - original draft preparation: JD, RS, GMSM, TJvH. Writing - review and editing: JD, RS, GMSM, MN, TJvH. Supervision: GMSM, TJvH.

All authors read, commented and approved the final manuscript.

## Acknowledgements

The authors would like to thank the patients and their family members for their collaboration. We thank Nicole van Koetsveld, Annemieke Trebitsch, Ayse Sener, Sally den Boer, Chantal Elling, Cindy Becht and Jamilieh Hosseini for culturing the fibroblasts. We thank Gideon Huigen, Roy Lamping, Martine van Amelsvoort, Lida Prins and Chérise Jurriens for performing the RNA isolations. Furthermore, we would like to thank Dr. Stefan Barakat for critically reading the manuscript.

## Notes

### Competing Interest Statement

The authors have declared no competing interest.

### Author Declarations

Ethics committee/IRB of Erasmus MC gave ethical approval for this work

