## Supplemental Figures 1-5 for "RNA-sequencing improves diagnosis for neurodevelopmental disorders by identifying pathogenic non-coding variants and reinterpretation of coding variants"

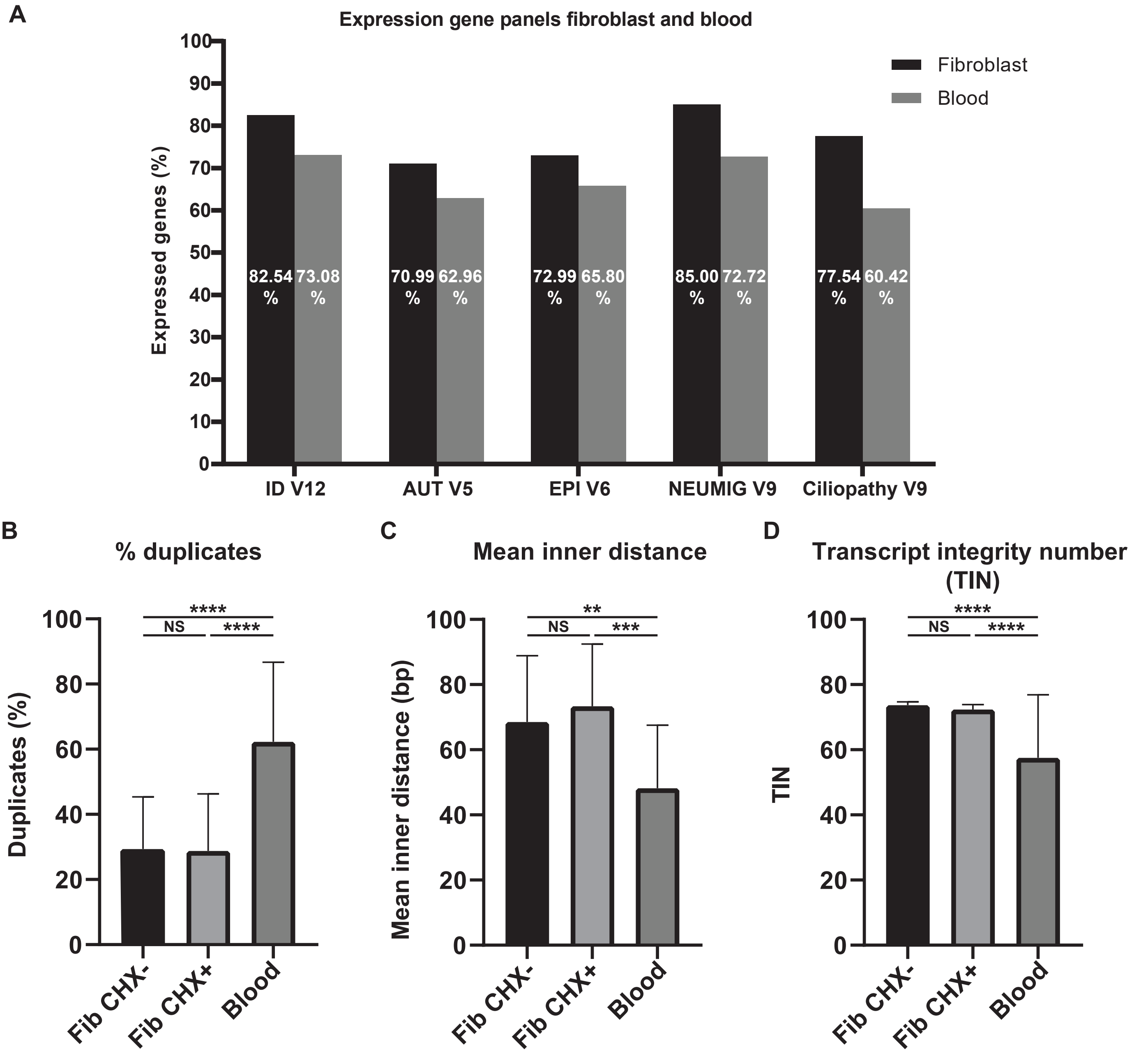


**Fig. S1.** Gene panel expression and quality control of fibroblast and PAX. **A** Percentages of genes that are expressed in each of five neurological gene panels for fibroblast and blood. A gene was considered expressed if the median across all fibroblast samples has at least 25 reads of the fragment in a total library size of 80 million reads (TPM≥ 0.3125). Number of genes in each panel: ID V12 = 1226, AUT V5 = 162, EPI V6 = 348, NEUMIG V9 = 219, Ciliopathy V9 = 187. Overlap of panels with ID V12 panel: AUT V5 = 145/162 (89.5%) , EPI V6 = 259/348 (74.4%), NEUMIG V9 = 194/219 (88.6%), Ciliopathy V9 = 40/187 (21.4%). **B, C, D** Quality control of RNA-seq data from fibroblasts with and without CHX, and PAXgene blood samples. PAX has a significantly decreased quality on % duplicates , mean inner distance and transcript integrity number compared with the fibroblast samples. Number of samples, blood = 10, Fibroblast CHX- = 47 Fibroblast CHX+ = 49. Statistical test: one-way ANOVA with Tukey's multiple comparisons test. *p<0.05, **p<0.01, ***p<0.001, ****p<0.0001

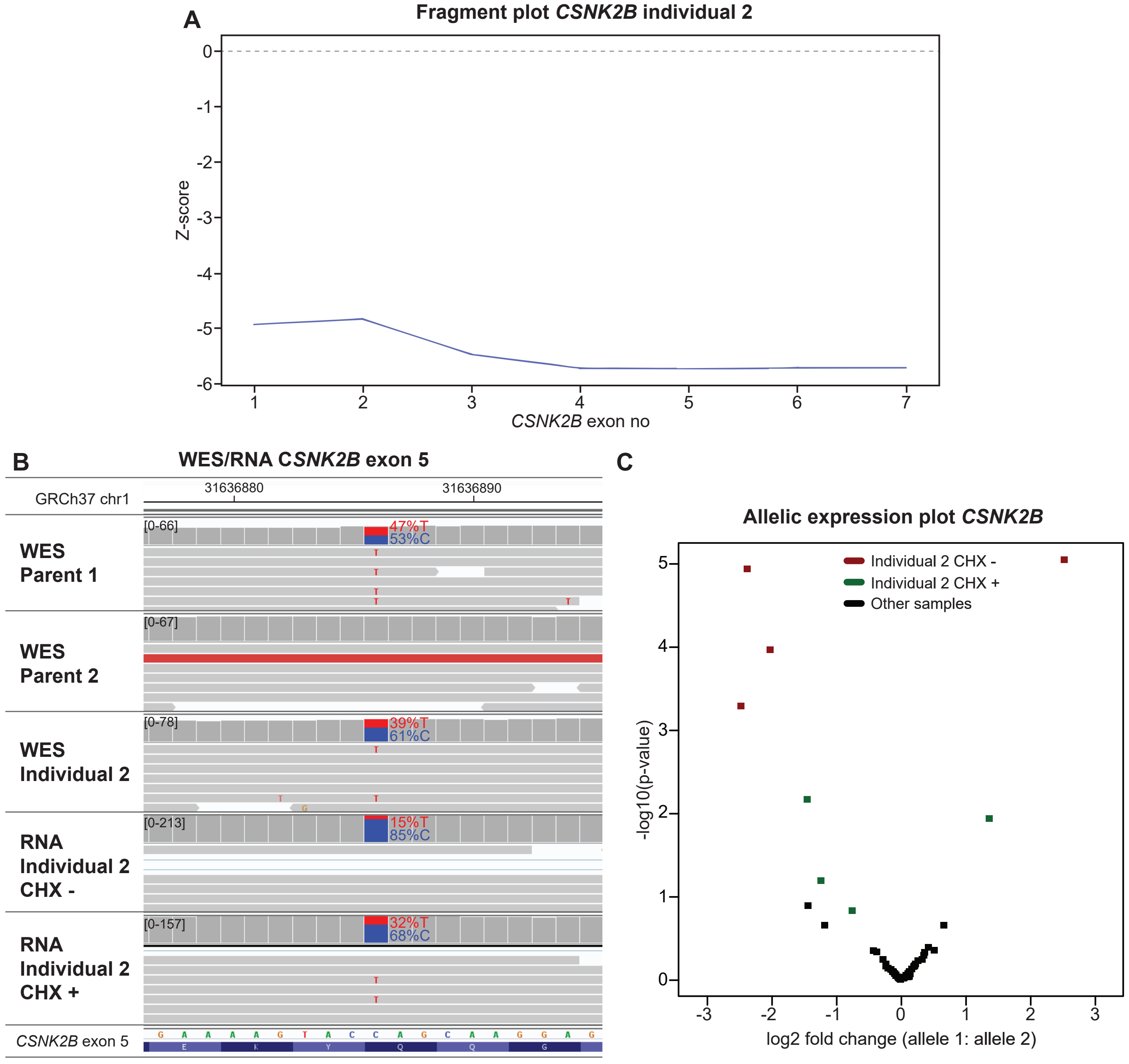


**Fig. S2.** Mono-allelic expression of CSNK2B. **A** Individual 2's fragment plot of CSNK2B reveals a downregulation of all exons of CSNK2B. **B** IGV plot of WES trio data (individual 2 and parents) and RNA-seq data (mapped reads) of untreated and CHX treated samples of individual 2 of CSNK2B exon 5 region. The stop-gain variant CSNK2B:c.304C>T, p.(Gln102*), was inherited from parent 1 and results in MAE. **C** Allelic expression plot of CSNK2B. Each dot shows the log2 fold change of one SNV calculated from the RNA-seq data reads. The four SNV with the highest allelic imbalance are from individual 2's untreated fibroblast (red).

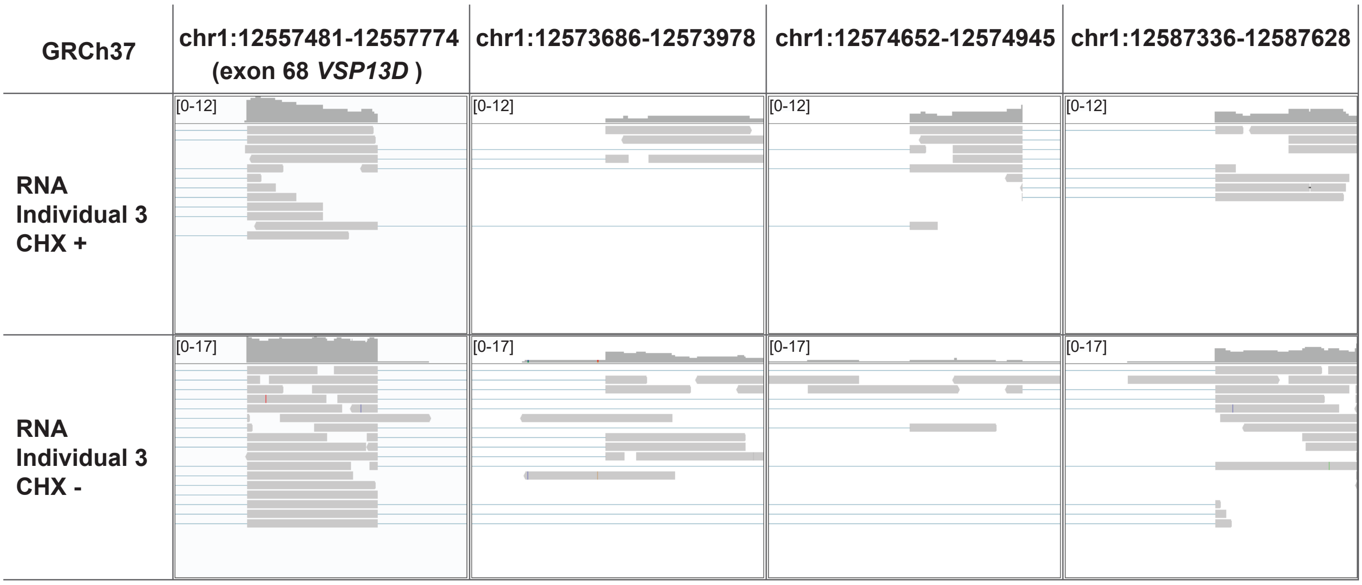


**Fig. S3.** Splicing from VPS13D exon 68 occurs to three different non-coding regions in individual 3. IGV plot of RNA-seq data (mapped reads) of untreated and CHX treated samples of individual 3 of VPS13D exon 68 and three genomic non-coding regions downstream of VPS13D. Reads mapping to exon 68 are splitted towards three other non-coding genomic regions downstream of VPS13D. the genomic locations of the splice acceptor sites are: Chr1(GRCh37):g.12573821, Chr1(GRCh37):g.12574795 and Chr1(GRCh37):g.12587487.

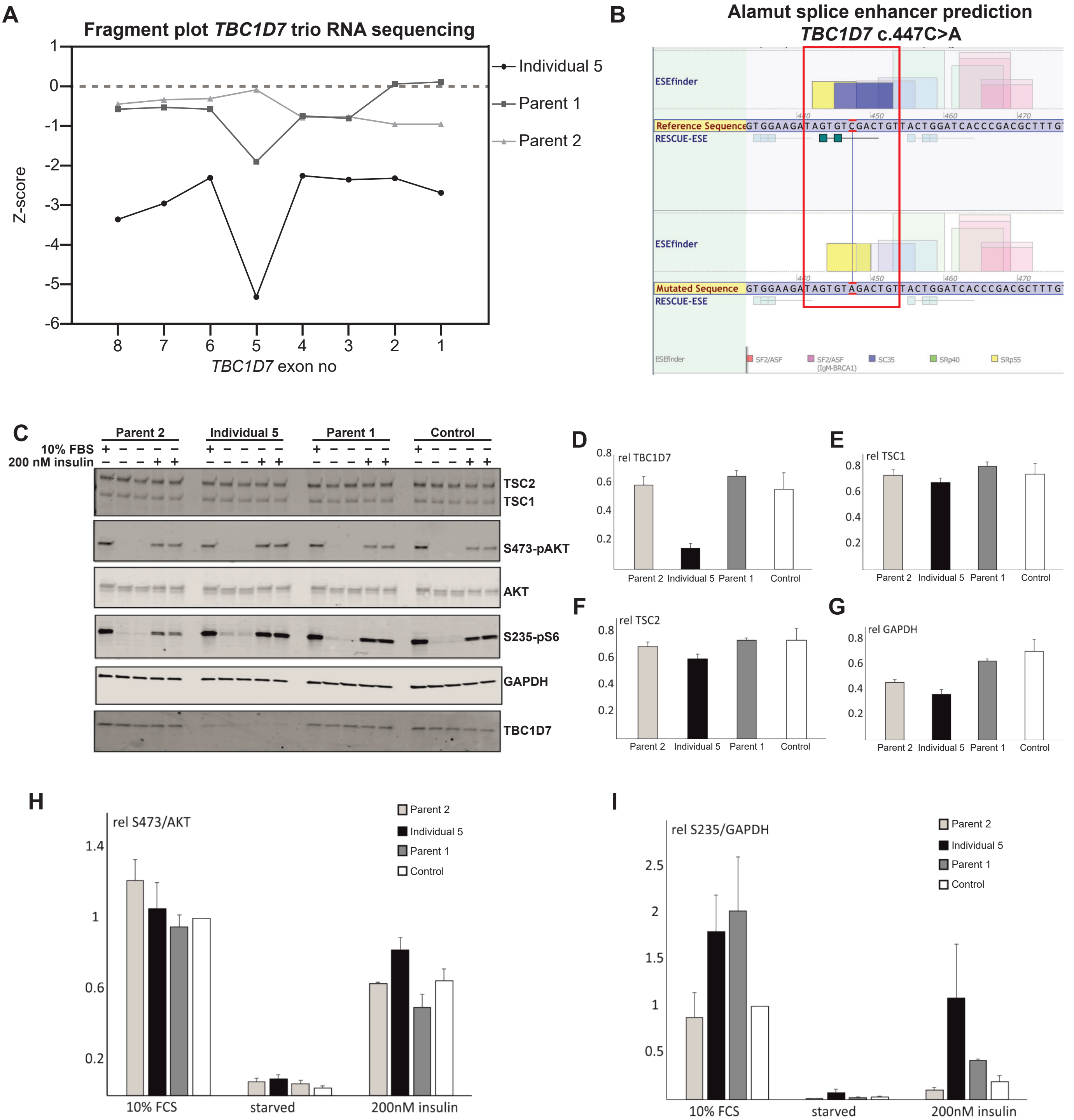


**Fig. S4.** TBC1D7 western blot and TSC-mTOR pathway analysis. **A** Fragment plot of TBC1D7 trio RNA sequencing (individual 5 and parents). Both parent 1 and individual 5 have a decreased expression of exon 5, where in individual 5 expression of all TBC1D7 exons is decreased. **B** Alamut splice enhancer prediction of TBC1D7:c.447C>A,p.Val149= shows a loss of SC35 splice enhancers site and two RESCUE-ESE sites. **C** Immuno-blot showing that individual 5's fibroblast have a decreased TBC1D7 abundance. **D, E, F, G** Intensities of the protein bands detected by immunoblotting c were quantified; n=3. D TBC1D7 abundance is decreased in the index , but not in the parents or an unrelated control. **H, I** Relative Ser473 phosphorylation of AKT (**H**) and Ser235/Ser236 phos-phorylation of ribosomal protein S6 (**I**) was estimated from the immunoblot quantification. No clear differences in AKT Ser473 or S6 Ser235/Ser236 phosphorylation were detected. Upon serum starvation, mTOR signaling was clearly down-regulated.

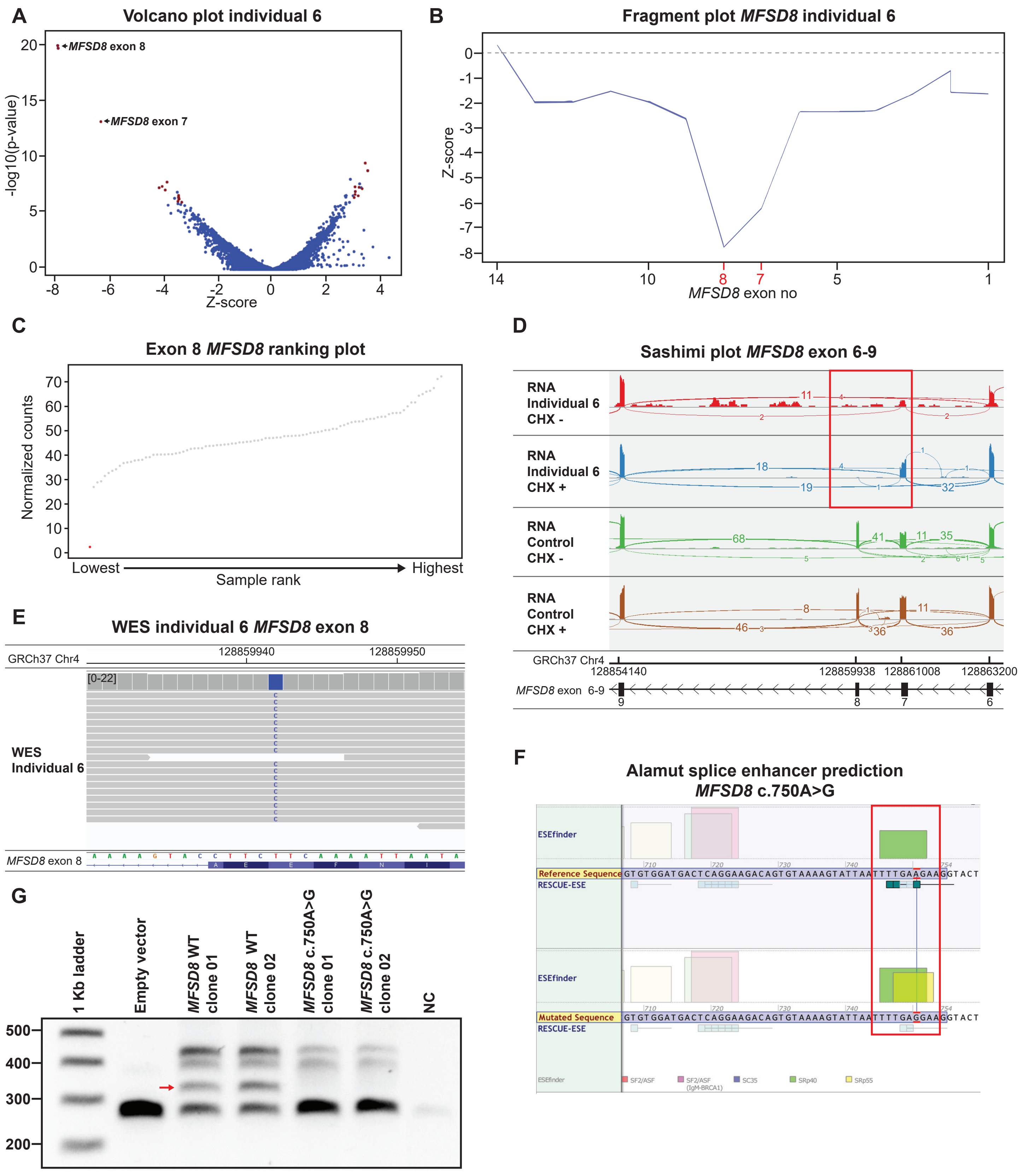


**Fig. S5.** Synonymous variant MFSD8 causes exon skipping. **A** Individual 6's Volcano plot (Z-threshold=4; p-value<0.0025; ID panel) indicating MFSD8 exon 7 and 8 are the most downregulated exons. The two most downregulated exons refer both to MFSD8 exon 8, though to two different transcripts (NM_001371590.1 and NM_001371595.1). **B** Individual 6's fragment plot of MFSD8 shows a downregulation of exon 7 and 8 MFSD8. **C** Exon ranking plot of exon 8 of MFSD8, individual 6 (red dot) has almost no counts, where on average in control samples ~40 counts are seen (grey dots). **D** Sashimi plot RNA-seq data (mapped reads) of untreated and CHX treated samples of individual 6 and a control of MFSD8 exon 6-9 showing skipping of exon 7 and 8 in individual 6 (red box). **E** Individual 6's WES data reveals a homozygous synonymous variant MFSD8:c.750A>G p.Glu250= in exon 8. **F** Splice enhancer prediction in Alamut showing loss of 3 RESCUE-ESE sites, but also a SRp55 ESE gain. **G** Minigene exon trapping assay confirms that the MFSD8 missense variant c.750A>G, p.(Glu250=) increases the likelihood of exon skipping. Red arrow indicates the RT-PCR product where MFSD8 exon 8 inclusion occurred.
